# Viral And Antibody Testing For Coronavirus Disease 2019 (Covid-19): Factors Associated With Positivity In Electronic Health Records From The United States

**DOI:** 10.1101/2021.03.19.21253924

**Authors:** Lisa Lindsay, Matthew H. Secrest, Shemra Rizzo, Dan Keebler, Fei Yang, Larry W. Tsai

## Abstract

**Background:** Insufficient information on SARS-CoV-2 testing results exists in clinical practice from the United States.

**Methods:** We conducted an observational retrospective cohort study using Optum® de-identified COVID-19 electronic health records from the United States to characterize patients who received a SARS-CoV-2 viral or antibody test between February 20, 2020 and July 10, 2020. We assessed temporal trends in testing and positivity by demographic and clinical characteristics; evaluated concordance between viral and antibody tests; and identified factors associated with positivity via multivariable logistic regression.

**Results:** Our study population included 891,754 patients. Overall positivity rate for SARS-CoV-2 was 9% and 12% for viral and antibody tests, respectively. Positivity rate was inversely associated with the number of individuals tested and decreased over time across regions and race/ethnicities. Among patients who received a viral test followed by an antibody test, concordance ranged from 90%-93% depending on the duration between the two tests which is notable given uncertainties related to specific viral and antibody test characteristics. The following factors increased the odds of viral and antibody positivity in multivariable models: male, Hispanic or non-Hispanic Black and Asian, uninsured or Medicaid insurance, Northeast residence, dementia, diabetes, and obesity. Charlson Comorbidity Index was negatively associated with test positivity. We identified symptoms that were positively associated with test positivity, as well as, commonly co-occurring symptoms / conditions. Pediatric patients had reduced odds of a positive viral test, but conversely had increased odds of a positive antibody test.

**Conclusions:** This study identified sociodemographic and clinical factors associated with SARS-CoV-2 testing and positivity within routine clinical practice from the United States.

## Background

The global coronavirus disease 2019 (COVID-19) pandemic has taken an enormous toll on human health and the economy. As of March 16, 2021, surveillance in the United States (US) estimated approximately 29.5 million cases and 535,000 deaths across the nation with one of the highest incidence rates in the world [1, 2].

Severe acute respiratory syndrome coronavirus 2 (SARS-CoV-2) testing is a critical but challenging component of both surveillance and treatment decisions. Variable test access, quality and reliability have been key considerations in our understanding of COVID-19 [3, 4, 5, 6, 7, 8]. While surveillance data and targeted studies provide important information related to patient characteristics and symptoms, a large-scale systematic assessment of factors associated with testing and positivity in clinical practice would provide important information pertaining to testing decisions and interpretation of viral and antibody tests [9, 10, 11, 12, 13, 14, 15, 16].

We sought to utilize electronic health records (EHRs) from the US to characterize patients tested for SARS-CoV-2 in routine clinical practice and to identify factors associated with positivity. We also sought to assess temporal trends in testing and positivity rates (PRs) and evaluate concordance between viral and antibody tests.

## METHODS

### Study Objectives

We aimed to 1) characterize patients tested for COVID-19 according to test type (viral, antibody) and result; 2) evaluate temporal trends in COVID-19 positivity rate (PR) by test type according to key factors; 3) assess concordance between viral and antibody test results among patients who received both types of tests, and 4) estimate the relative risk of a positive COVID-19 test result compared to a negative test result by test type accounting for important cofactors via logistic regression.

### Data Source

We used the Optum de-identified COVID-19 EHR dataset, containing patient-level health records from January 1, 2007 through July 10, 2020, to identify patients tested for SARS-CoV-2 or COVID-19 antibodies. This dataset was sourced from Optum’s longitudinal EHR repository derived from a national sample of inpatient and outpatient medical records from more than 700 hospitals and 7000 clinics. The COVID-19 EHR dataset included patients who had documented clinical care with a diagnosis of COVID-19 or acute respiratory illness after February 1, 2020 and/or COVID-19 testing (positive or negative result). Data was de-identified in compliance with the Health Insurance Portability and Accountability Act (HIPAA) Expert Determination Method and managed according to Optum customer data use agreements. The EHRs did not capture detailed information on the specific sub-type or manufacturer for COVID-19 viral and antibody tests that would have enabled assessment of test sensitivity and/or specificity. Antibody tests were available in the EHRs beginning the week of April 22, 2020. Further, the EHRs did not capture detailed information related to residence, such as zip code or independent versus institutional setting (e.g., nursing home).

### Study Population

We included patients of all ages, if they satisfied the criteria required for entry into one or more of the following testing cohorts and the index date occurred between February 20, 2020 and July 10, 2020. We did not require patients to have a minimum follow-up period.

#### Cohort Definitions

Cohort 1 (C1) consisted of patients with a laboratory test or procedural code for either a viral (molecular or antigen test) or antibody testing based on Logical Observation Identifiers Names and Codes (LOINC), Current Procedural Terminology (CPT), Healthcare Common Procedure Coding System (HCPC), Proprietary Laboratory Analyses (PLA) codes or string search for test names (Supplement A). We considered the C1 index date as the first COVID-19 testing, regardless of type (viral or antibody).

As a subset of C1, we defined Cohort 2 (C2) as patients whose first observed COVID-19 testing was a viral test.

As a subset of C1, we indexed Cohort 3 (C3) patients on the first antibody test received based on laboratory or procedural data source as previously described. We identified C3 patients regardless of prior viral test to enable full characterization of antibody testing and assessment of concordance between viral and antibody tests.

### Data Analysis

For C2 and C3 patients, we classified test results as positive, negative, and not definitive.

Clinical context/setting variables, including symptoms and vital signs (0-7 days pre-index) were defined using diagnostic codes (Supplement B). We chose a 0-7 day window to ensure that identified symptoms were related to the reason for testing.

Additionally, we adapted a definition of time since first COVID-like illness (CLI) based on the CLI definition endorsed by the Centers for Disease Control and Prevention (CDC) & Council of State and Territorial Epidemiologists [17] (Supplement C). We identified comorbidities using diagnostic codes (≥1 code in any position 0-365 days pre-index; Supplement D). We utilized string searches and LOINC codes to evaluate laboratory tests for respiratory isolates (0-7 days pre/post-index).

### Descriptive Analysis

We evaluated the distribution of variables and PRs by cohort and definite test result. To assess the representativeness of our study population, we compared the distribution of key sociodemographic factors in our study cohorts to the all Optum beneficiaries, and the 2019 US census population.

We evaluated temporal trends in several variables (including PR). We utilized an UpSet plot to assess co-occurring symptoms / conditions among viral positive patients. We assessed concordance between test results for patients who received definitive viral and antibody results.

### Models to Identify Factors Impacting Test Positivity

Multivariable logistic regression models were fit to assess factors impacting SARS-CoV-2 positivity for the following binary outcomes: a) positive initial viral test among C2 patients and b) positive initial antibody test among C3 patients.

Models included patients who had a definite test result and ≥365 days pre-index database history to ensure capture of existing comorbidities. Candidate variables for the models included those identified from prior research and commonly (>3% of patients) occurring symptoms. Non-significant interactions and collinear variables were not included. In both models, multiple imputation was used to address missing data for variables with <25% of missingness using multivariate imputation by chained equations (Supplement E and F).

The final models included sociodemographics, clinical/setting variables, comorbidities, region, and study week. Additionally, the C2 model included a variable for co-infections and an interaction term between region and study week. While the C3 model included prior viral test results.

## RESULTS

### Descriptive

We identified 891,754 patients in our overall study population (n=806,510 in C2 with a first viral test; n=95,930 in C3 with an antibody test at any time; n=15,182 in both C2 and C3; n=4,496 in C1 but not in C2 or C3 due to unknown test type) (Figure 1). These patients were predominantly adult, female, non-Hispanic Whites, commercially insured, and Midwest/Northeast residents (Table 1). A majority of patients had no CLI identified nor underlying comorbidity (Supplement G and H). Pediatric patients were underrepresented in our study population relative to both the Optum and census populations.

**Figure 1.**
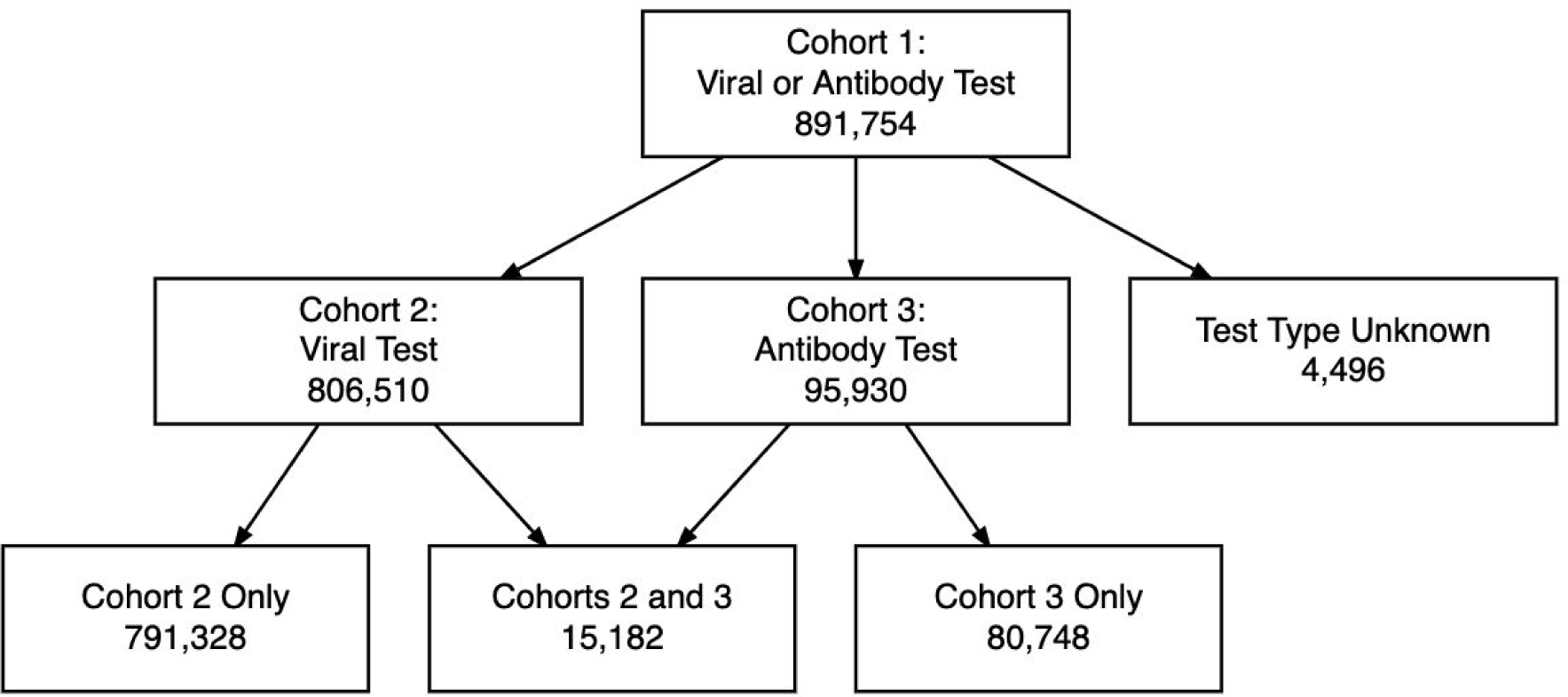
Flow Diagram of Study Population

**Table 1.**
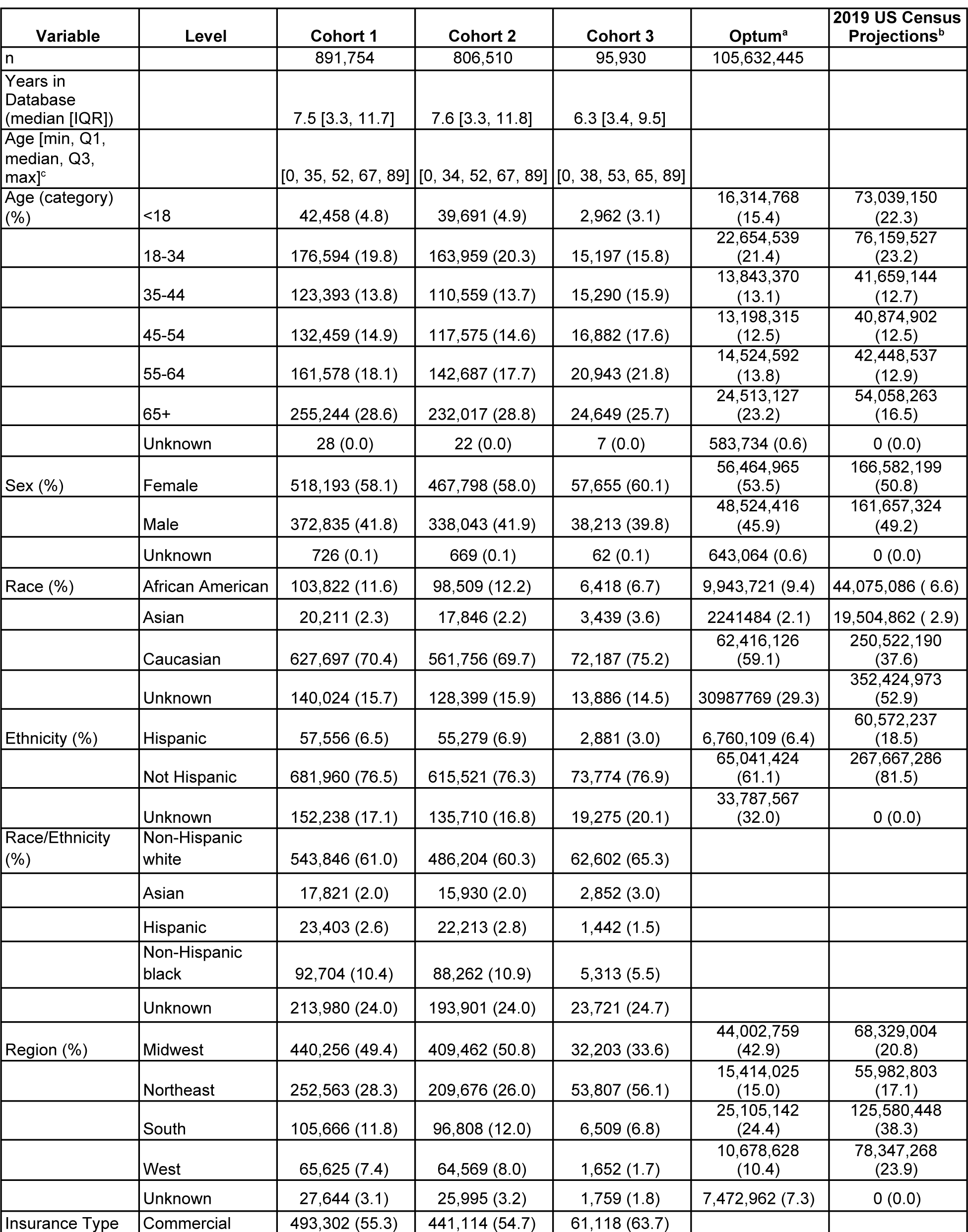
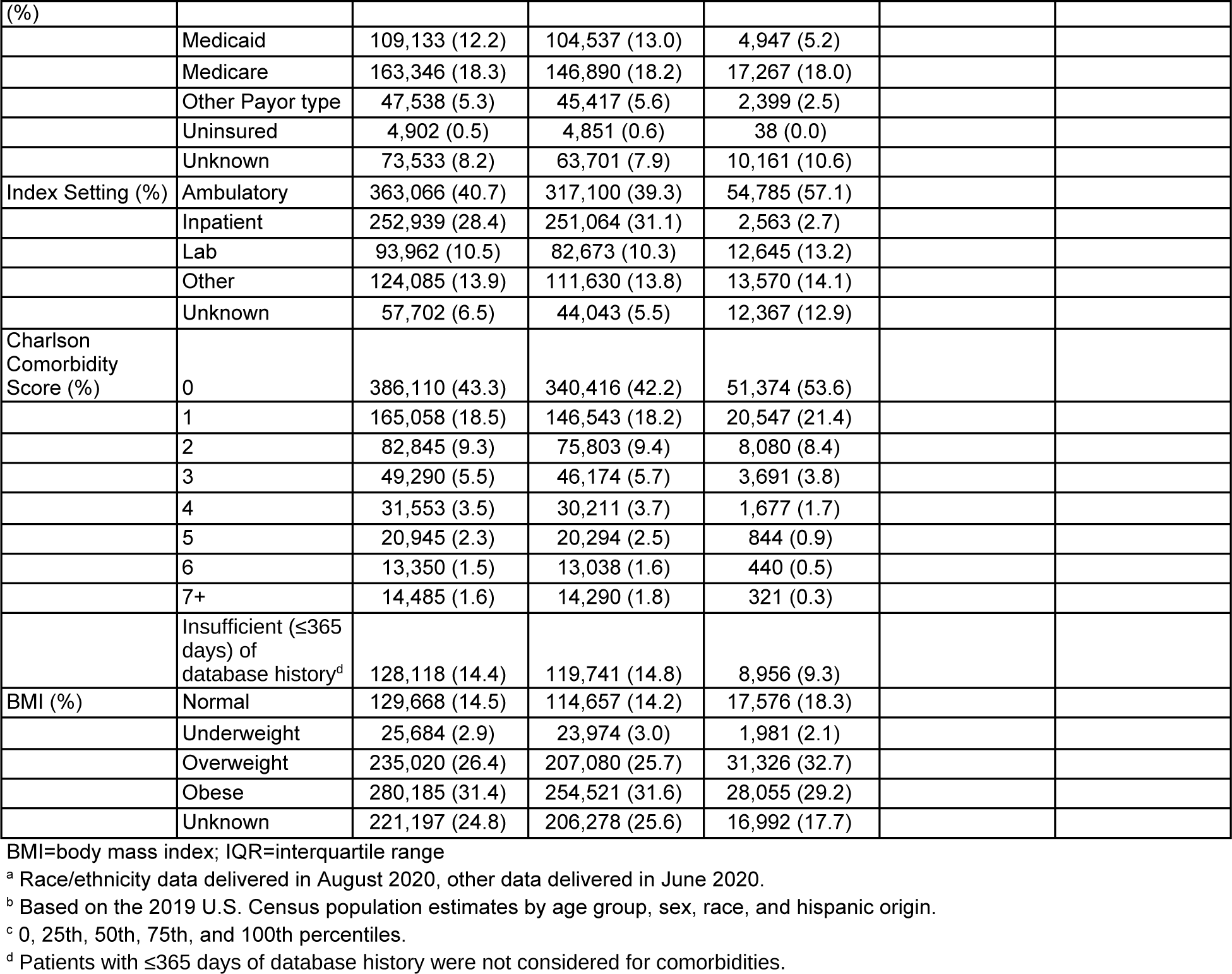
Description of Study Population (Cohorts 1-3) and Comparison to Source Population(s)

### Temporal Trends

SARS-CoV-2 viral test reporting increased steadily in the week of March 11, 2020 with PRs peaking 2 weeks later driven primarily by data from the Northeast. Antibody test reporting began the week of April 22, 2020 with a concurrent modest peak in PRs, again driven largely by Northeast data (Figure 2). There was an inverse relationship between testing and positivity over time and most regions. Positivity rate was inversely associated with the number of individuals tested and decreased over time across regions and race/ethnicities (Figure 2).

**Figure 2.**
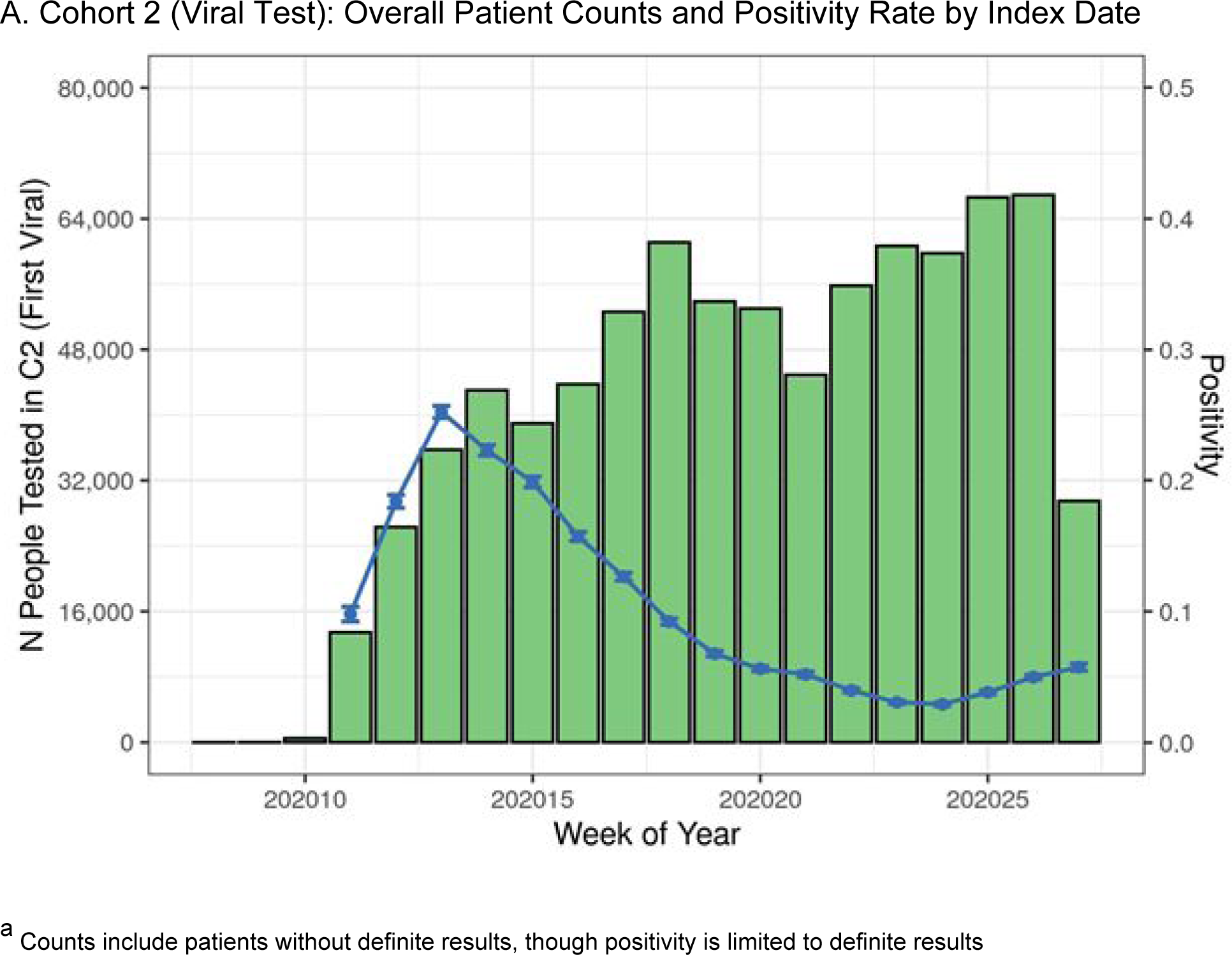
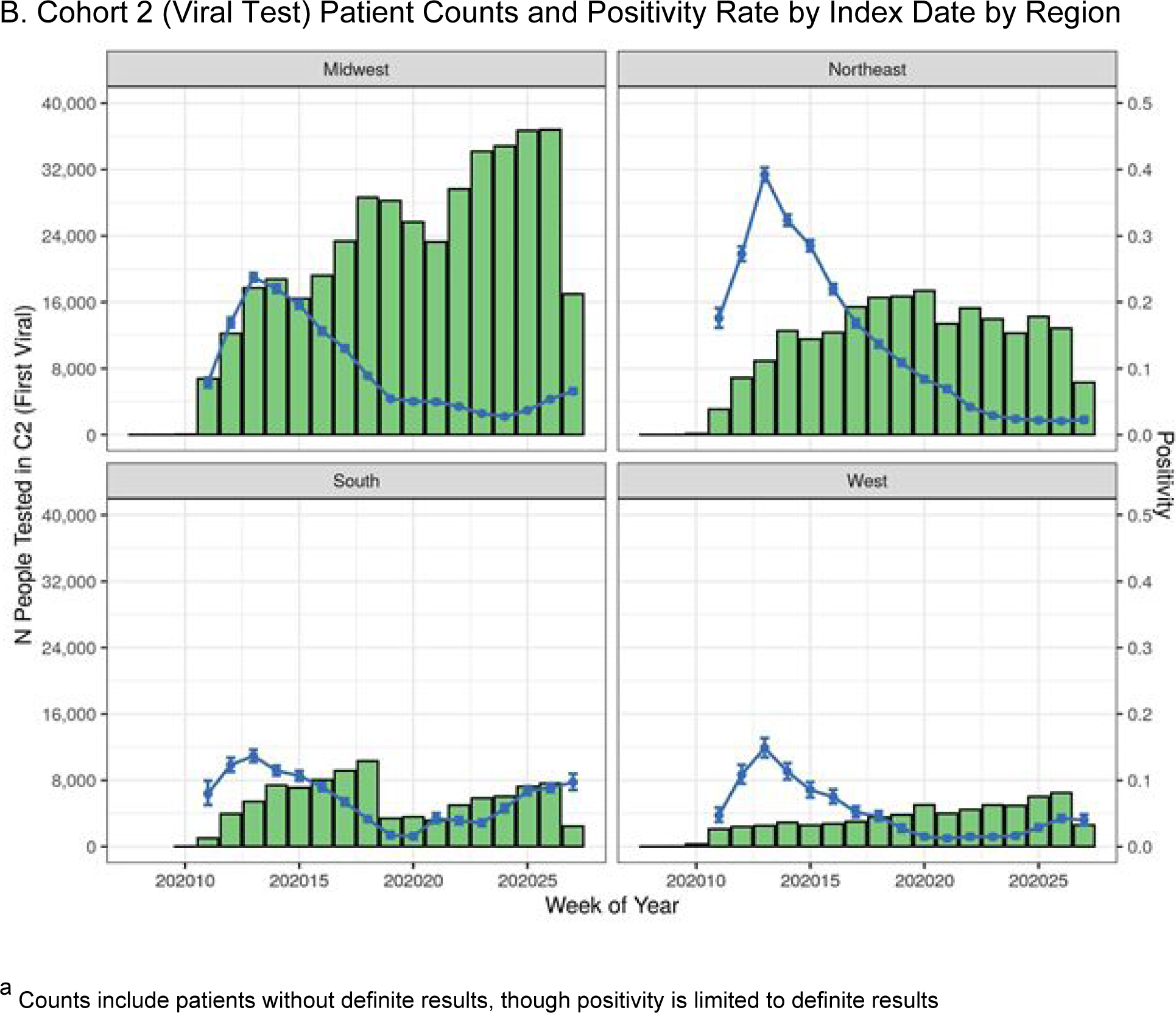
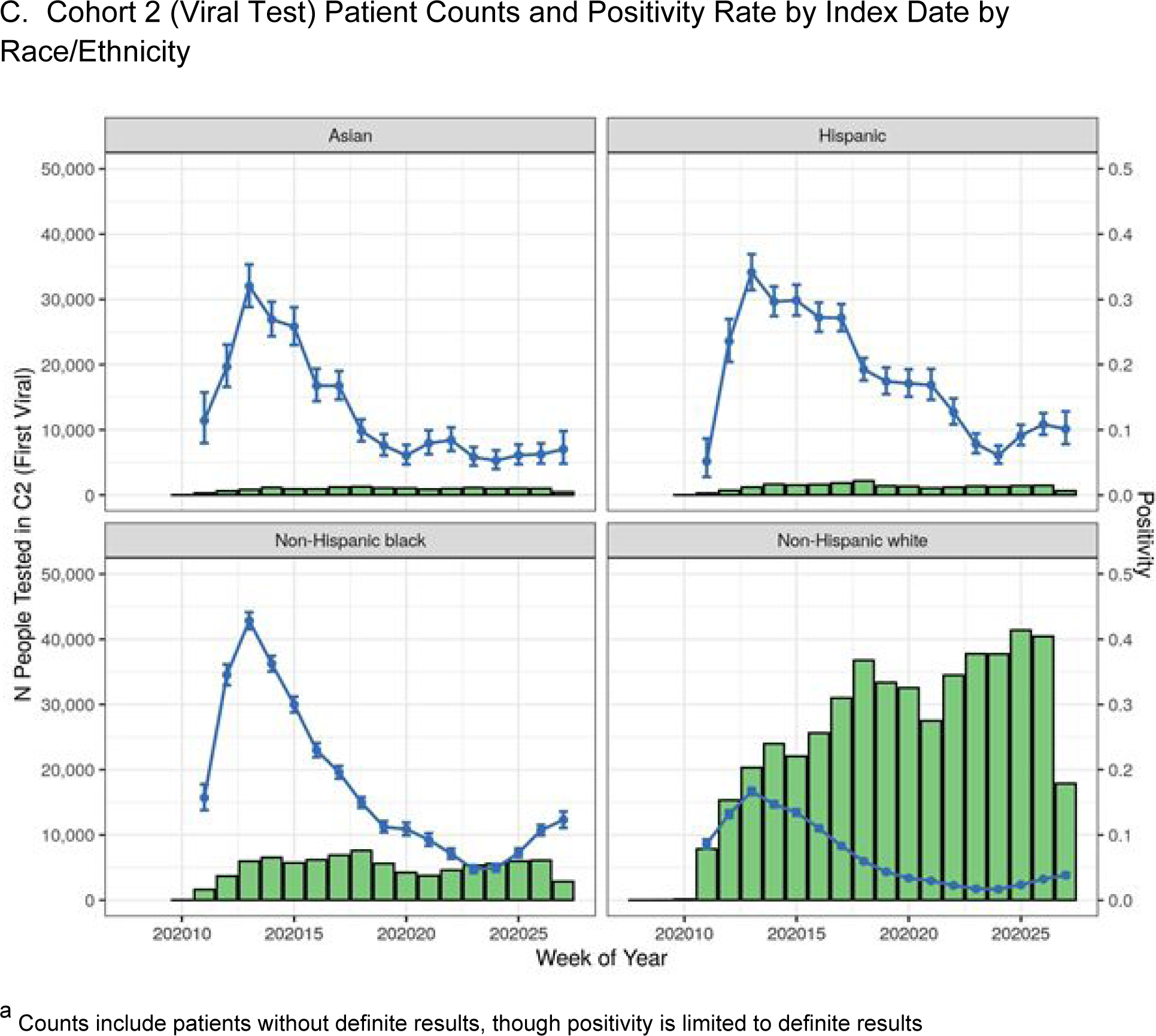
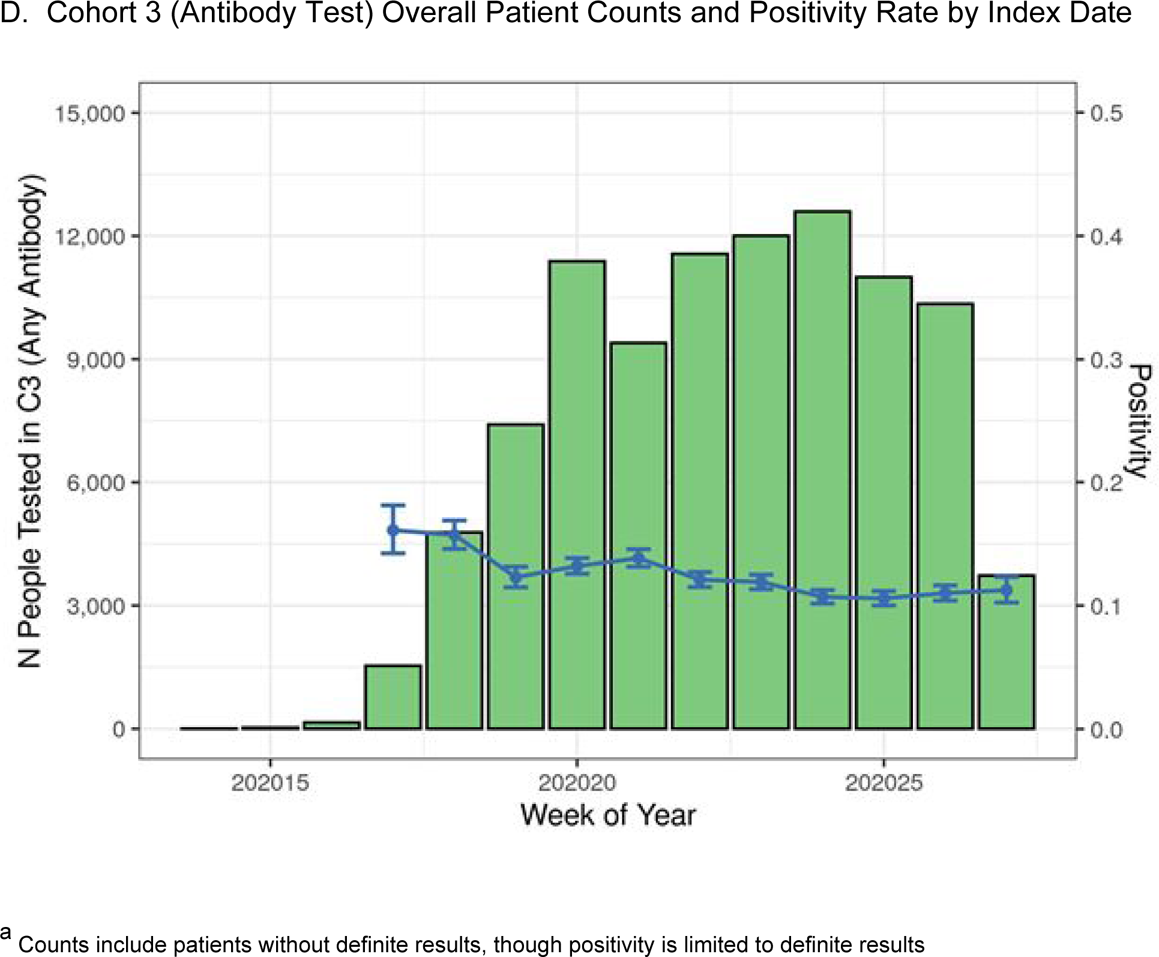
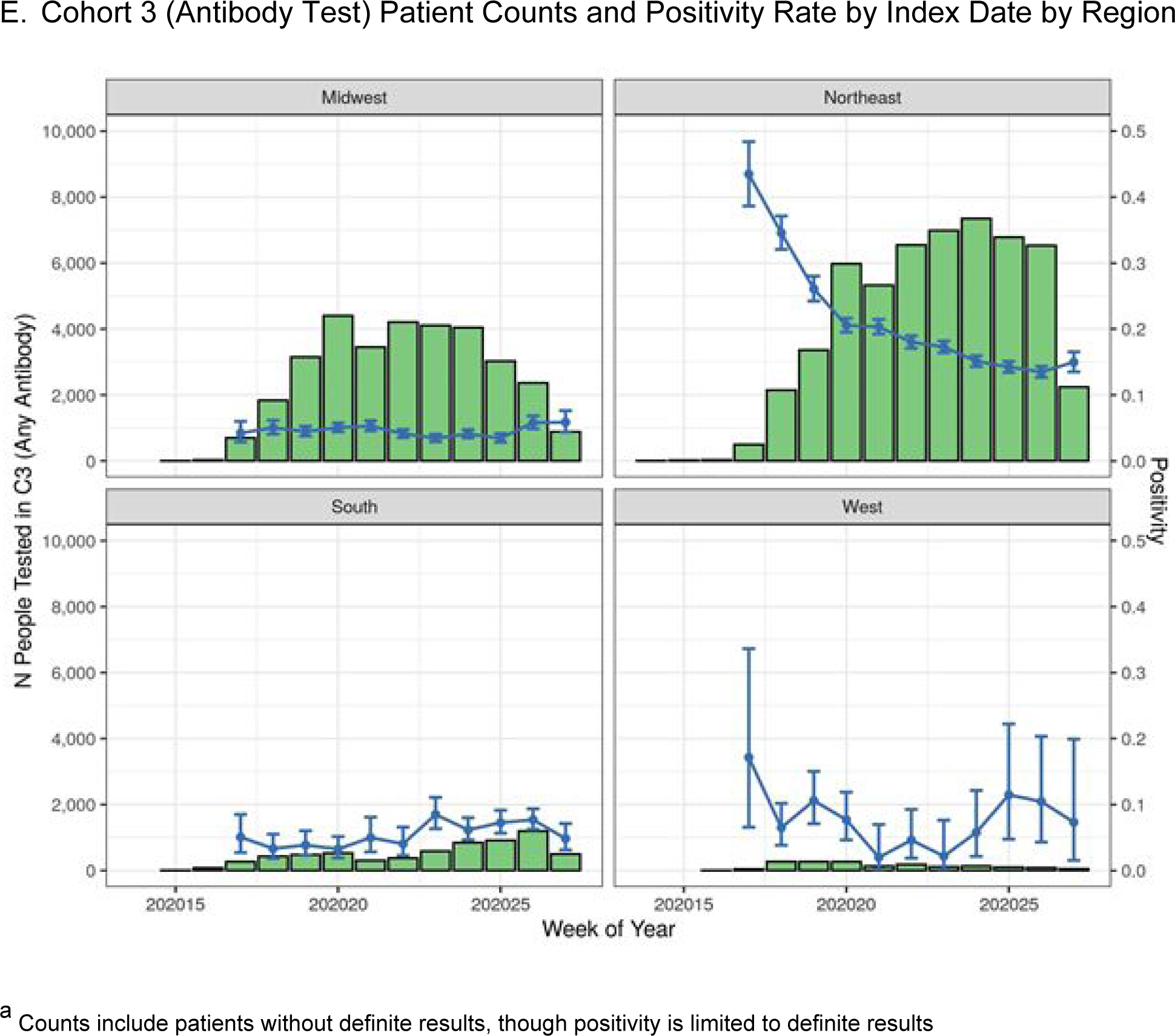
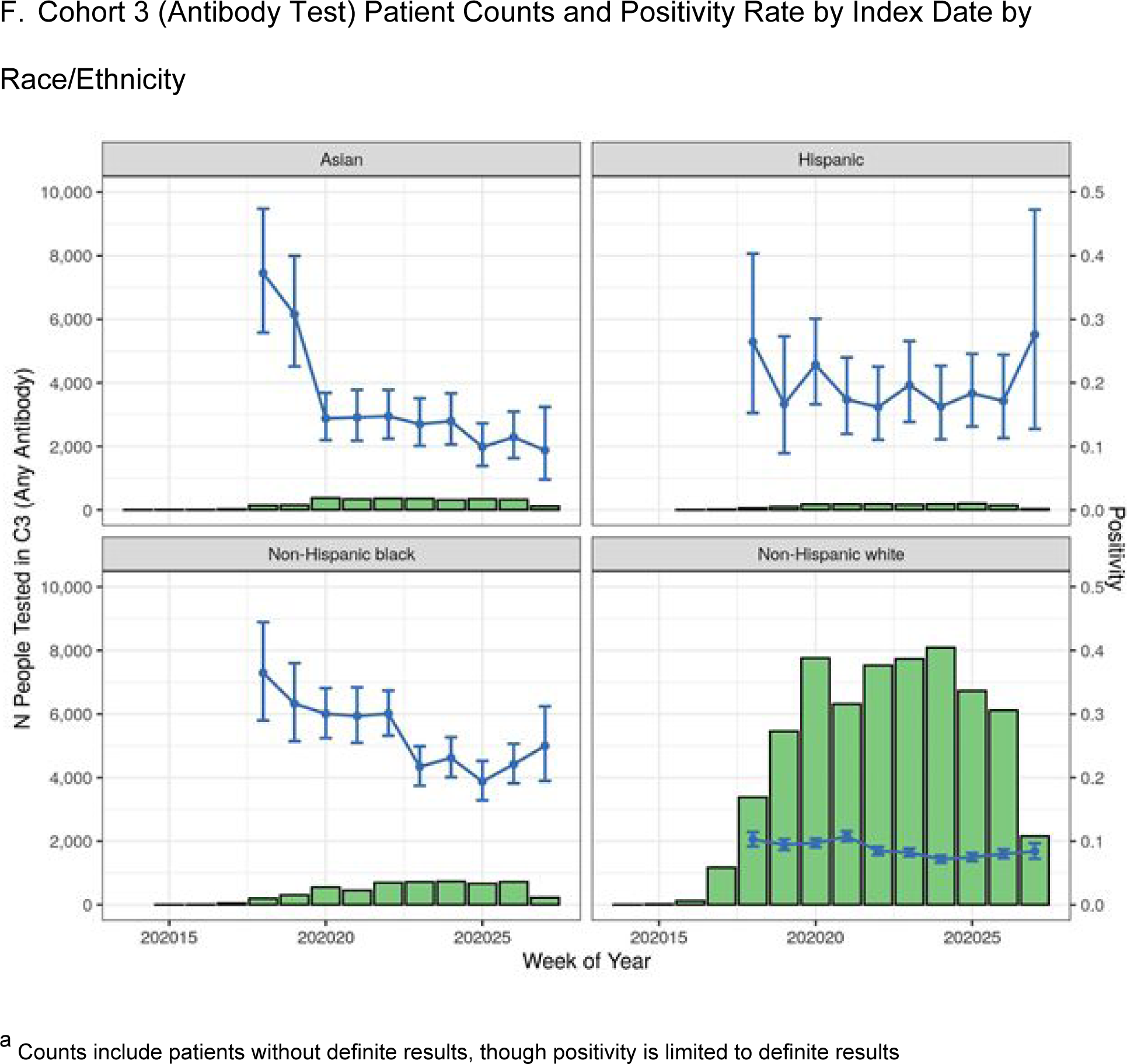
Patient Count and Positivity by Cohort, Index Date, Region, and Race/Ethnicity (Figures A-F)a

### Cohort 2

Among 806,510 patients in C2, 771,278 (96%) had definite results.

C2 consisted largely of adult (95%), female (58%), non-Hispanic (77%), White (70%), residents of the Midwest (51%), and those who had commercial insurance (55%; Table 2). Approximately 50% of patients with >1 year of database history had a Charlson Comorbidity Index (CCI) = 0 compared to approximately 40% of US adults [18], while 77% of our study population (with non-missing body mass index (BMI)) were overweight or obese compared to approximately 72% of US adults [19]. Information on the type of specimen collected for a viral test was missing in 81% of patients.

**Table 2.** Cohort 2: SARS-CoV-2 Positivity Rates by Sociodemographic and Clinical Characteristics

| Variable | Level | Positivity (%) | Test Result |  |  |  |  |
| --- | --- | --- | --- | --- | --- | --- | --- |
|  |  |  | Overall | Positive | Negative | Uninterpretable | None<sup>a</sup> |
| n |  | 9.1 | 806,510 | 70,472 | 700,806 | 11,002 | 24,230 |
| Age [min, Q1, median, Q3, max]<sup>b</sup> |  | NA | [0, 34, 52, 67, 89] | [0, 34, 51, 65, 89] | [0, 34, 52, 67, 89] | [0, 36, 55, 68, 89] | [0, 32, 50, 65, 89] |
| Age (category) (%) | <18 | 5.7 | 39,691 (4.9) | 2,120 (3.0) | 35,121 (5.0) | 580 (5.3) | 1,870 (7.7) |
|  | 18-34 | 9.9 | 163,959 (20.3) | 15,600 (22.1) | 141,405 (20.2) | 1,973 (17.9) | 4,981 (20.6) |
|  | 35-44 | 9.6 | 110,559 (13.7) | 10,150 (14.4) | 95,703 (13.7) | 1,353 (12.3) | 3,353 (13.8) |
|  | 45-54 | 10.3 | 117,575 (14.6) | 11,602 (16.5) | 100,770 (14.4) | 1,578 (14.3) | 3,625 (15.0) |
|  | 55-64 | 9.1 | 142,687 (17.7) | 12,444 (17.7) | 123,915 (17.7) | 2,000 (18.2) | 4,328 (17.9) |
|  | 65+ | 8.3 | 232,017 (28.8) | 18,556 (26.3) | 203,883 (29.1) | 3,518 (32.0) | 6,060 (25.0) |
|  | Unknown | 0.0 | 22 (0.0) | 0 (0.0) | 9 (0.0) | 0 (0.0) | 13 (0.1) |
| Sex (%) | Female | 8.5 | 467,798 (58.0) | 38,171 (54.2) | 409,203 (58.4) | 6,401 (58.2) | 14,023 (57.9) |
|  | Male | 10.0 | 338,043 (41.9) | 32,262 (45.8) | 291,036 (41.5) | 4,587 (41.7) | 10,158 (41.9) |
|  | Unknown | 6.4 | 669 (0.1) | 39 (0.1) | 567 (0.1) | 14 (0.1) | 49 (0.2) |
| Race (%) | African American | 17.9 | 98,509 (12.2) | 16,912 (24.0) | 77,435 (11.0) | 1,209 (11.0) | 2,953 (12.2) |
|  | Asian | 12.7 | 17,846 (2.2) | 2,227 (3.2) | 15,254 (2.2) | 87 (0.8) | 278 (1.1) |
|  | Caucasian | 6.3 | 561,756 (69.7) | 34,030 (48.3) | 502,014 (71.6) | 8,576 (77.9) | 17,136 (70.7) |
|  | Other/Unknown | 14.0 | 128,399 (15.9) | 17,303 (24.6) | 106,103 (15.1) | 1,130 (10.3) | 3,863 (15.9) |
| Ethnicity (%) | Hispanic | 22.3 | 55,279 (6.9) | 11,877 (16.9) | 41,365 (5.9) | 652 (5.9) | 1,385 (5.7) |
|  | Not Hispanic | 8.1 | 615,521 (76.3) | 47,523 (67.4) | 541,895 (77.3) | 9,336 (84.9) | 16,767 (69.2) |
|  | Unknown | 8.6 | 135,710 (16.8) | 11,072 (15.7) | 117,546 (16.8) | 1,014 (9.2) | 6,078 (25.1) |
| Race/Ethnicity (%) | Non-Hispanic white | 5.8 | 486,204 (60.3) | 27,158 (38.5) | 437,551 (62.4) | 7,927 (72.1) | 13,568 (56.0) |
|  | Asian | 12.7 | 15,930 (2.0) | 1,990 (2.8) | 13,618 (1.9) | 75 (0.7) | 247 (1.0) |
|  | Hispanic | 19.2 | 22,213 (2.8) | 4,084 (5.8) | 17,217 (2.5) | 250 (2.3) | 662 (2.7) |
|  | Non-Hispanic black | 18 | 88,262 (10.9) | 15,231 (21.6) | 69,478 (9.9) | 1,107 (10.1) | 2,446 (10.1) |
|  | Unknown | 11.9 | 193,901 (24.0) | 22,009 (31.2) | 162,942 (23.3) | 1,643 (14.9) | 7,307 (30.2) |
| Region (%) | Midwest | 8.4 | 409,462 (50.8) | 32,556 (46.2) | 352,918 (50.4) | 7,132 (64.8) | 16,856 (69.6) |
|  | Northeast | 12.7 | 209,676 (26.0) | 26,144 (37.1) | 179,041 (25.5) | 1,578 (14.3) | 2,913 (12.0) |
|  | South | 7.4 | 96,808 (12.0) | 6,991 (9.9) | 87,054 (12.4) | 889 (8.1) | 1,874 (7.7) |
|  | West | 4.3 | 64,569 (8.0) | 2,629 (3.7) | 59,047 (8.4) | 1,043 (9.5) | 1,850 (7.6) |
|  | Unknown | 8.6 | 25,995 (3.2) | 2,152 (3.1) | 22,746 (3.2) | 360 (3.3) | 737 (3.0) |
| Insurance Type (%) | Commercial | 9.1 | 441,114 (54.7) | 38,451 (54.6) | 383,626 (54.7) | 5,729 (52.1) | 13,308 (54.9) |
|  | Medicaid | 10.3 | 104,537 (13.0) | 10,251 (14.5) | 89,236 (12.7) | 1,288 (11.7) | 3,762 (15.5) |
|  | Medicare | 7.6 | 146,890 (18.2) | 10,774 (15.3) | 130,425 (18.6) | 2,017 (18.3) | 3,674 (15.2) |
|  | Other Payor type | 11.0 | 45,417 (5.6) | 4,720 (6.7) | 38,317 (5.5) | 1,098 (10.0) | 1,282 (5.3) |
|  | Uninsured | 16.4 | 4,851 (0.6) | 657 (0.9) | 3,347 (0.5) | 317 (2.9) | 530 (2.2) |
|  | Unknown | 9.1 | 63,701 (7.9) | 5,619 (8.0) | 55,855 (8.0) | 553 (5.0) | 1,674 (6.9) |
| Index Setting (%) | Ambulatory | 6.9 | 317,100 (39.3) | 21,069 (29.9) | 282,486 (40.3) | 1,889 (17.2) | 11,656 (48.1) |
|  | Inpatient | 12.2 | 251,064 (31.1) | 29,273 (41.5) | 211,064 (30.1) | 2,984 (27.1) | 7,743 (32.0) |
|  | Lab | 6.7 | 82,673 (10.3) | 5,130 (7.3) | 71,205 (10.2) | 5,156 (46.9) | 1,182 (4.9) |
|  | Other | 10.8 | 111,630 (13.8) | 11,793 (16.7) | 97,206 (13.9) | 716 (6.5) | 1,915 (7.9) |
|  | Unknown | 7.6 | 44,043 (5.5) | 3,207 (4.6) | 38,845 (5.5) | 257 (2.3) | 1,734 (7.2) |
| Test source (%) | Upper respiratory | 8.6 | 154,995 (19.2) | 13,033 (18.5) | 139,125 (19.9) | 2,837 (25.8) | 0 (0.0) |
|  | Lower respiratory | 11.5 | 26 (0.0) | 3 (0.0) | 23 (0.0) | 0 (0.0) | 0 (0.0) |
|  | Unknown | 9.3 | 651,489 (80.8) | 57,436 (81.5) | 561,658 (80.1) | 8,165 (74.2) | 24230 (100.0) |
| Positive for any seasonal respiratory isolate<sup>c</sup> (%) | At least 1 test positive | 6.9 | 267 (0.0) | 17 (0.0) | 231 (0.0) | 9 (0.1) | 10 (0.0) |
|  | Not tested | 8.2 | 722,101 (89.5) | 56,756 (80.5) | 635,904 (90.7) | 8,363 (76.0) | 21,078 (87.0) |
|  | Tested but no positives | 17.5 | 84,142 (10.4) | 13,699 (19.4) | 64,671 (9.2) | 2,630 (23.9) | 3,142 (13.0) |
| Days since COVID-like illness (%) | No COVID-like illness observed | 6.8 | 614,872 (76.2) | 39,786 (56.5) | 547,006 (78.1) | 8,654 (78.7) | 19,426 (80.2) |
|  | 0 | 19.5 | 125,608 (15.6) | 23,609 (33.5) | 97,291 (13.9) | 1,500 (13.6) | 3,208 (13.2) |
|  | 1 - 3 | 15.4 | 25,845 (3.2) | 3,844 (5.5) | 21,192 (3.0) | 289 (2.6) | 520 (2.1) |
|  | 4 - 7 | 12.7 | 7,319 (0.9) | 893 (1.3) | 6,162 (0.9) | 94 (0.9) | 170 (0.7) |
|  | 8 - 14 | 9.6 | 7,169 (0.9) | 666 (0.9) | 6,255 (0.9) | 107 (1.0) | 141 (0.6) |
|  | 15 - 21 | 7.5 | 5,756 (0.7) | 412 (0.6) | 5,117 (0.7) | 71 (0.6) | 156 (0.6) |
|  | 22 - 28 | 6.3 | 5,832 (0.7) | 349 (0.5) | 5,216 (0.7) | 99 (0.9) | 168 (0.7) |
|  | 29 - 35 | 6.8 | 6,061 (0.8) | 396 (0.6) | 5,401 (0.8) | 73 (0.7) | 191 (0.8) |
|  | 36 - 42 | 6.7 | 8,048 (1.0) | 517 (0.7) | 7,166 (1.0) | 115 (1.0) | 250 (1.0) |
| Number of symptoms<sup>d</sup> (%) | 0 | 4.7 | 455,212 (56.4) | 20,208 (28.7) | 414,359 (59.1) | 6,676 (60.7) | 13,969 (57.7) |
|  | 1-3 | 12.6 | 321,632 (39.9) | 38,906 (55.2) | 269,426 (38.4) | 3,712 (33.7) | 9,588 (39.6) |
|  | 4-7 | 39.4 | 29,290 (3.6) | 11,049 (15.7) | 16,960 (2.4) | 610 (5.5) | 671 (2.8) |
|  | 8-11 | 83.5 | 376 (0.0) | 309 (0.4) | 61 (0.0) | 4 (0.0) | 2 (0.0) |
|  | 12+ | NaN | 0 (0.0) | 0 (0.0) | 0 (0.0) | 0 (0.0) | 0 (0.0) |
| Charlson Comorbidity Score (%) | 0 | 9.5 | 340,416 (42.2) | 30,822 (43.7) | 295,097 (42.1) | 4,760 (43.3) | 9,737 (40.2) |
|  | 1 | 8.4 | 146,543 (18.2) | 11,784 (16.7) | 127,850 (18.2) | 2,067 (18.8) | 4,842 (20.0) |
|  | 2 | 7.8 | 75,803 (9.4) | 5,659 (8.0) | 66,827 (9.5) | 1,055 (9.6) | 2,262 (9.3) |
|  | 3 | 8.0 | 46,174 (5.7) | 3,548 (5.0) | 40,656 (5.8) | 633 (5.8) | 1,337 (5.5) |
|  | 4 | 8.0 | 30,211 (3.7) | 2,311 (3.3) | 26,733 (3.8) | 381 (3.5) | 786 (3.2) |
|  | 5 | 7.8 | 20,294 (2.5) | 1,527 (2.2) | 17,951 (2.6) | 326 (3.0) | 490 (2.0) |
|  | 6 | 7.4 | 13,038 (1.6) | 936 (1.3) | 11,642 (1.7) | 188 (1.7) | 272 (1.1) |
|  | 7+ | 7.1 | 14,290 (1.8) | 984 (1.4) | 12,785 (1.8) | 212 (1.9) | 309 (1.3) |
|  | Insufficient (≤365) days of database history<sup>e</sup> | 11.3 | 119,741 (14.8) | 12,901 (18.3) | 101,265 (14.4) | 1,380 (12.5) | 4,195 (17.3) |
| BMI (%) | Normal | 6.8 | 114,657 (14.2) | 7,513 (10.7) | 102,790 (14.7) | 1,382 (12.6) | 2,972 (12.3) |
|  | Underweight | 5.0 | 23,974 (3.0) | 1,137 (1.6) | 21,554 (3.1) | 332 (3.0) | 951 (3.9) |
|  | Overweight | 8.4 | 207,080 (25.7) | 16,718 (23.7) | 182,420 (26.0) | 2,698 (24.5) | 5,244 (21.6) |
|  | Obese | 9.1 | 254,521 (31.6) | 22,155 (31.4) | 221,506 (31.6) | 3,788 (34.4) | 7,072 (29.2) |
|  | Unknown | 11.7 | 206,278 (25.6) | 22,949 (32.6) | 172,536 (24.6) | 2,802 (25.5) | 7,991 (33.0) |

BMI=body mass index; IQR=interquartile range
<sup>a</sup> Indexed on procedural code without corresponding laboratory result
<sup>b</sup> 0, 25th, 50th, 75th, and 100th percentiles
<sup>c</sup> influenza, adenovirus, non-SARS-
CoV-2 coronavirus, human metapneumovirus, human rhinovirus/enterovirus, parainfluenza virus, respiratory syncytial virus, chlamydomphila pneumonia, mycoplasma pneumonia, and/or bordetella pertussis
<sup>d</sup> Symptoms were chosen as those associated with positivity in C2 modeling
<sup>e</sup> Patients with ≤365 days of database history were not considered for comorbidities

The overall PR in C2 was 9%. PR was slightly higher among patients tested in the hospital (12%) and among those without insurance (16%).

Figure 3 displays common symptoms and intersecting sets of these symptoms in patients who had a positive viral test in C2.

**Figure 3.**
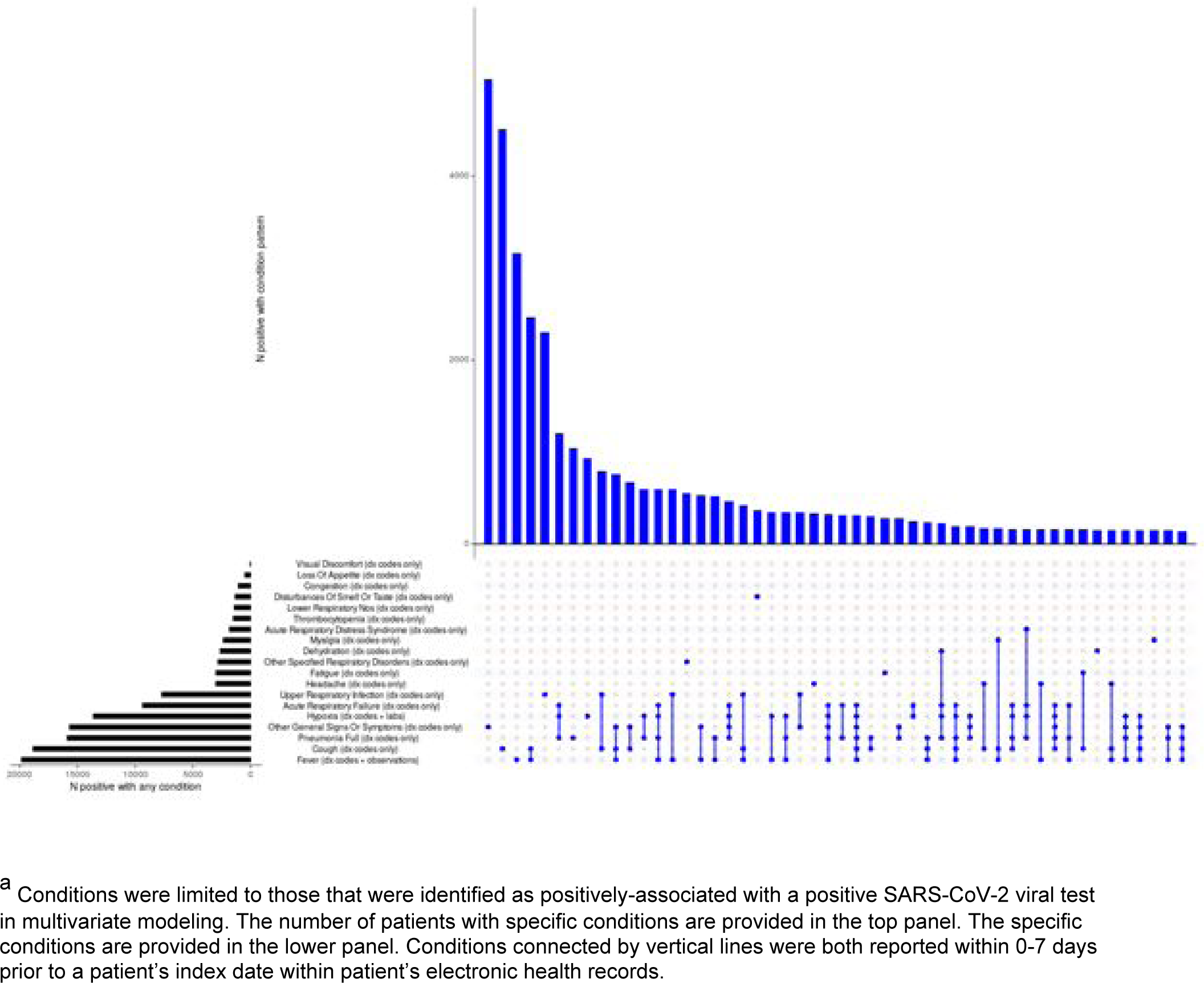
Frequency and Co-Occurrence of Select Conditions in Patients Positive for SARS-CoV-2 via Viral Test

Most C2 patients did not have a CLI recorded in the 6 weeks prior to their test (76% of C2 patients and 57% viral positive patients; Figure 4; Table 2). Among those with a prior CLI, the median interval between reported CLI and initial definitive test was 0 days and 83% occurred ≤7 days of their test result. Coinfections were rare (<0.1%).

**Figure 4.**
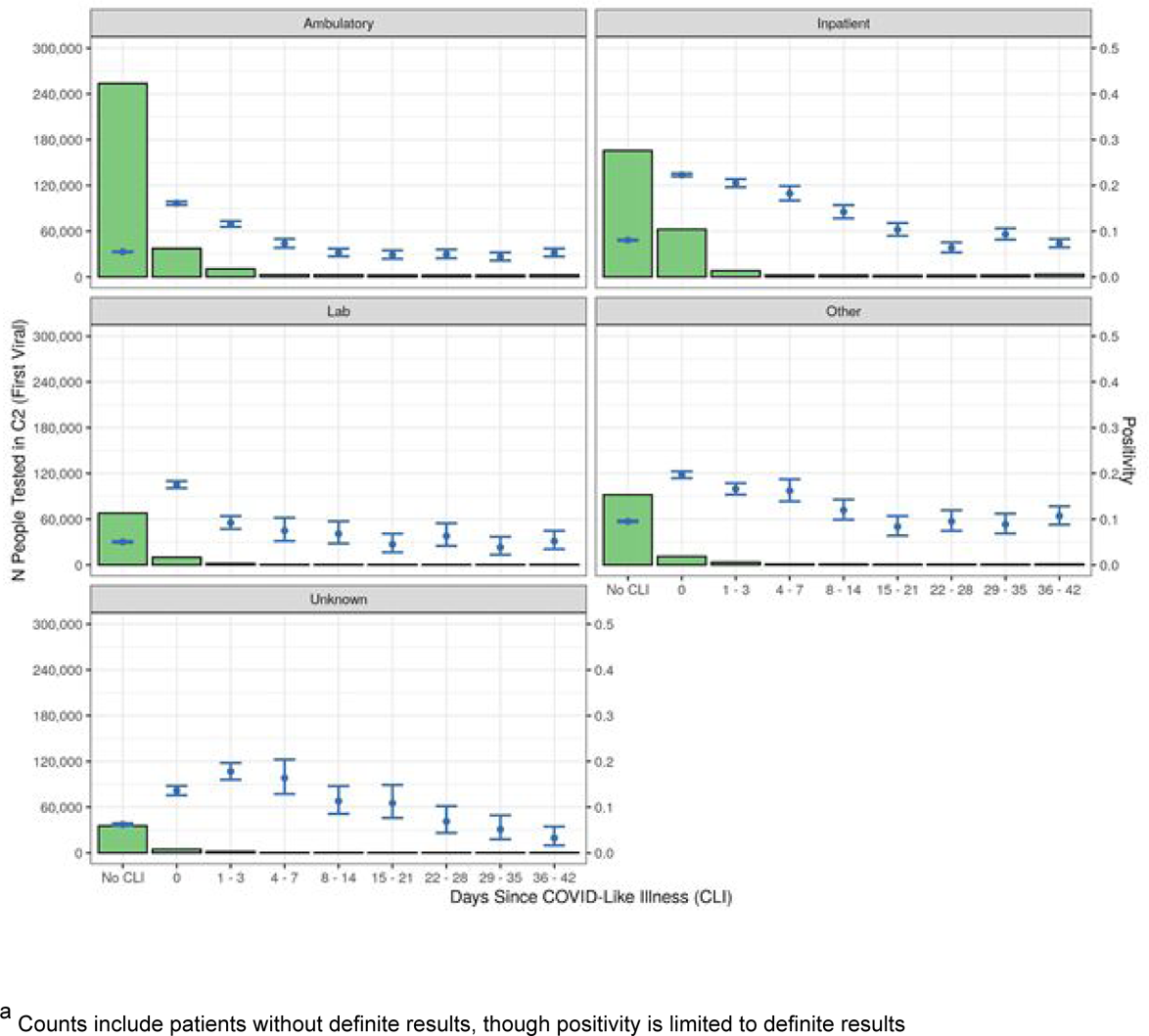
SARS-CoV-2 Viral Test Positivity and Days Since COVID-like Illness by Setting ^a^

### Cohort 3

Among 95,930 patients in C3, 91,741 (96%) had definite results.

Similar to C2, C3 patients were predominantly adult (97%), female (60%), non-Hispanic (77%), White (75%), and commercially insured (64%; Table 3). Slightly more than half of the patients lived in the Northeast (56% vs. 26% for C2). Approximately 75% of patients (with a reported BMI) were overweight or obese and 59% of patients with >1 year of database history had a CCI score=0.

**Table 3.** Cohort 3: SARS-CoV-2 Positivity Rates by Sociodemographic and Clinical Characteristics

| Variable | Level | Positivity (%) | Test Result |  |  |  |  |
| --- | --- | --- | --- | --- | --- | --- | --- |
|  |  |  | Overall | Positive | Negative | Uninterpretable | None<sup>a</sup> |
| n |  | 12.1 | 95,930 | 11,094 | 80,647 | 2,438 | 1,751 |
| Age [min, Q1, median, Q3, max]<sup>b</sup> |  | NA | [0, 38, 53, 65, 89] | [0, 37, 53, 64, 89] | [0, 38, 53, 65, 89] | [2, 35, 47, 60, 89] | [0, 40, 55, 65, 89] |
| Age (category) (%) | <18 | 17.4 | 2,962 (3.1) | 490 (4.4) | 2,331 (2.9) | 35 (1.4) | 106 (6.1) |
|  | 18-34 | 13.0 | 15,197 (15.8) | 1,880 (16.9) | 12,585 (15.6) | 529 (21.7) | 203 (11.6) |
|  | 35-44 | 10.5 | 15,290 (15.9) | 1,527 (13.8) | 12,949 (16.1) | 578 (23.7) | 236 (13.5) |
|  | 45-54 | 12.8 | 16,882 (17.6) | 2,065 (18.6) | 14,109 (17.5) | 405 (16.6) | 303 (17.3) |
|  | 55-64 | 12.1 | 20,943 (21.8) | 2,417 (21.8) | 17,621 (21.8) | 471 (19.3) | 434 (24.8) |
|  | 65+ | 11.4 | 24,649 (25.7) | 2,715 (24.5) | 21,051 (26.1) | 420 (17.2) | 463 (26.4) |
|  | Unknown | 0.0 | 7 (0.0) | 0 (0.0) | 1 (0.0) | 0 (0.0) | 6 (0.3) |
| Sex (%) | Female | 11.5 | 57,655 (60.1) | 6,328 (57.0) | 48,662 (60.3) | 1,605 (65.8) | 1,060 (60.5) |
|  | Male | 13.0 | 38,213 (39.8) | 4,762 (42.9) | 31,934 (39.6) | 833 (34.2) | 684 (39.1) |
|  | Unknown | 7.3 | 62 (0.1) | 4 (0.0) | 51 (0.1) | 0 (0.0) | 7 (0.4) |
| Race (%) | African American | 25.4 | 6,418 (6.7) | 1,571 (14.2) | 4,618 (5.7) | 145 (5.9) | 84 (4.8) |
|  | Asian | 15.5 | 3,439 (3.6) | 507 (4.6) | 2,769 (3.4) | 135 (5.5) | 28 (1.6) |
|  | Caucasian | 9.5 | 72,187 (75.2) | 6,555 (59.1) | 62,542 (77.6) | 1,642 (67.4) | 1,448 (82.7) |
|  | Other/Unknown | 18.7 | 13,886 (14.5) | 2,461 (22.2) | 10,718 (13.3) | 516 (21.2) | 191 (10.9) |
| Ethnicity (%) | Hispanic | 20.9 | 2,881 (3.0) | 552 (5.0) | 2,089 (2.6) | 98 (4.0) | 142 (8.1) |
|  | Not Hispanic | 10.5 | 73,774 (76.9) | 7,398 (66.7) | 63,270 (78.5) | 1,744 (71.5) | 1,362 (77.8) |
|  | Unknown | 17.1 | 19,275 (20.1) | 3,144 (28.3) | 15,288 (19.0) | 596 (24.4) | 247 (14.1) |
| Race/Ethnicity (%) | Non-Hispanic white | 8.7 | 62,602 (65.3) | 5,227 (47.1) | 54,742 (67.9) | 1,396 (57.3) | 1,237 (70.6) |
|  | Asian | 15.0 | 2,852 (3.0) | 406 (3.7) | 2,297 (2.8) | 123 (5.0) | 26 (1.5) |
|  | Hispanic | 18.9 | 1,442 (1.5) | 253 (2.3) | 1,083 (1.3) | 39 (1.6) | 67 (3.8) |
|  | Non-Hispanic black | 25.7 | 5,313 (5.5) | 1,315 (11.9) | 3,810 (4.7) | 121 (5.0) | 67 (3.8) |
|  | Unknown | 17.2 | 23,721 (24.7) | 3,893 (35.1) | 18,715 (23.2) | 759 (31.1) | 354 (20.2) |
| Region (%) | Midwest | 4.5 | 32,203 (33.6) | 1,416 (12.8) | 30,133 (37.4) | 52 (2.1) | 602 (34.4) |
|  | Northeast | 17.8 | 53,807 (56.1) | 9,030 (81.4) | 41,637 (51.6) | 2,151 (88.2) | 989 (56.5) |
|  | South | 5.9 | 6,509 (6.8) | 380 (3.4) | 6,050 (7.5) | 41 (1.7) | 38 (2.2) |
|  | West | 7.2 | 1,652 (1.7) | 102 (0.9) | 1,305 (1.6) | 156 (6.4) | 89 (5.1) |
|  | Unknown | 9.8 | 1,759 (1.8) | 166 (1.5) | 1,522 (1.9) | 38 (1.6) | 33 (1.9) |
| Insurance Type (%) | Commercial | 11.7 | 61,118 (63.7) | 6,820 (61.5) | 51,510 (63.9) | 1,473 (60.4) | 1,315 (75.1) |
|  | Medicaid | 24.7 | 4,947 (5.2) | 1,165 (10.5) | 3,556 (4.4) | 91 (3.7) | 135 (7.7) |
|  | Medicare | 12.0 | 17,267 (18.0) | 2,019 (18.2) | 14,744 (18.3) | 292 (12.0) | 212 (12.1) |
|  | Other Payor type | 11.7 | 2,399 (2.5) | 273 (2.5) | 2,066 (2.6) | 24 (1.0) | 36 (2.1) |
|  | Uninsured | 9.7 | 38 (0.0) | 3 (0.0) | 28 (0.0) | 0 (0.0) | 7 (0.4) |
|  | Unknown | 8.5 | 10,161 (10.6) | 814 (7.3) | 8,743 (10.8) | 558 (22.9) | 46 (2.6) |
| Index Setting (%) | Ambulatory | 11.1 | 54,785 (57.1) | 5,920 (53.4) | 47,299 (58.6) | 381 (15.6) | 1,185 (67.7) |
|  | Inpatient | 20.6 | 2,563 (2.7) | 480 (4.3) | 1,851 (2.3) | 37 (1.5) | 195 (11.1) |
|  | Lab | 11.8 | 12,645 (13.2) | 1,456 (13.1) | 10,887 (13.5) | 99 (4.1) | 203 (11.6) |
|  | Other | 12.2 | 13,570 (14.1) | 1,629 (14.7) | 11,696 (14.5) | 190 (7.8) | 55 (3.1) |
|  | Unknown | 15.3 | 12,367 (12.9) | 1,609 (14.5) | 8,914 (11.1) | 1,731 (71.0) | 113 (6.5) |
| Test source)<br>(%) | Plasma | 100.0 | 55 (0.1) | 2 (0.0) | 0 (0.0) | 53 (2.2) | 0 (0.0) |
|  | Unknown | 12.1 | 95,875 (99.9) | 11092 (100.0) | 80647 (100.0) | 2,385 (97.8) | 1751 (100.0) |
| Positive for<br>any seasonal<br>respiratory<br>isolate (%) | At least 1 test<br>positive | 25.0 | 4 (0.0) | 1 (0.0) | 3 (0.0) | 0 (0.0) | 0 (0.0) |
|  | Not tested | 12.1 | 95,302 (99.3) | 11,012 (99.3) | 80,185 (99.4) | 2,418 (99.2) | 1,687 (96.3) |
|  | Tested but no<br>positives | 15.0 | 624 (0.7) | 81 (0.7) | 459 (0.6) | 20 (0.8) | 64 (3.7) |
| Days since<br>COVID-like<br>illness (%) | No COVID-like<br>illness observed | 9.0 | 79,264 (82.6) | 6,831 (61.6) | 69,307 (85.9) | 1,896 (77.8) | 1,230 (70.2) |
|  | 0 | 18.7 | 5,879 (6.1) | 1,038 (9.4) | 4,513 (5.6) | 158 (6.5) | 170 (9.7) |
|  | 1 - 3 | 22.8 | 2,307 (2.4) | 490 (4.4) | 1,660 (2.1) | 74 (3.0) | 83 (4.7) |
|  | 4 - 7 | 22.7 | 1,373 (1.4) | 295 (2.7) | 1,005 (1.2) | 30 (1.2) | 43 (2.5) |
|  | 8 - 14 | 24.9 | 1,347 (1.4) | 312 (2.8) | 942 (1.2) | 39 (1.6) | 54 (3.1) |
|  | 15 - 21 | 25.8 | 1,059 (1.1) | 255 (2.3) | 733 (0.9) | 39 (1.6) | 32 (1.8) |
|  | 22 - 28 | 34.0 | 1,052 (1.1) | 330 (3.0) | 642 (0.8) | 49 (2.0) | 31 (1.8) |
|  | 29 - 35 | 42.7 | 1,489 (1.6) | 583 (5.3) | 783 (1.0) | 71 (2.9) | 52 (3.0) |
|  | 36 - 42 | 47.5 | 2,160 (2.3) | 960 (8.7) | 1,062 (1.3) | 82 (3.4) | 56 (3.2) |
| Charlson<br>Comorbidity<br>Score (%) | 0 | 11.3 | 51,374 (53.6) | 5,528 (49.8) | 43,506 (53.9) | 1,522 (62.4) | 818 (46.7) |
|  | 1 | 12.4 | 20,547 (21.4) | 2,457 (22.1) | 17,293 (21.4) | 390 (16.0) | 407 (23.2) |
|  | 2 | 12.2 | 8,080 (8.4) | 947 (8.5) | 6,787 (8.4) | 168 (6.9) | 178 (10.2) |
|  | 3 | 13.6 | 3,691 (3.8) | 484 (4.4) | 3,064 (3.8) | 57 (2.3) | 86 (4.9) |
|  | 4 | 16.8 | 1,677 (1.7) | 268 (2.4) | 1,332 (1.7) | 26 (1.1) | 51 (2.9) |
|  | 5 | 15.1 | 844 (0.9) | 121 (1.1) | 681 (0.8) | 10 (0.4) | 32 (1.8) |
|  | 6 | 16.1 | 440 (0.5) | 67 (0.6) | 348 (0.4) | 11 (0.5) | 14 (0.8) |
|  | 7+ | 20.3 | 321 (0.3) | 61 (0.5) | 240 (0.3) | 5 (0.2) | 15 (0.9) |
|  | Insufficient<br>(≤365) days of<br>database<br>history<sup>c</sup> | 13.6 | 8,956 (9.3) | 1,161 (10.5) | 7,396 (9.2) | 249 (10.2) | 150 (8.6) |
| BMI (%) | Normal | 11.0 | 17,576 (18.3) | 1,840 (16.6) | 14,960 (18.5) | 510 (20.9) | 266 (15.2) |
|  | Underweight | 12.5 | 1,981 (2.1) | 235 (2.1) | 1,640 (2.0) | 49 (2.0) | 57 (3.3) |
|  | Overweight | 12.3 | 31,326 (32.7) | 3,702 (33.4) | 26,324 (32.6) | 780 (32.0) | 520 (29.7) |
|  | Obese | 13.3 | 28,055 (29.2) | 3,596 (32.4) | 23,354 (29.0) | 526 (21.6) | 579 (33.1) |
|  | Unknown | 10.7 | 16,992 (17.7) | 1,721 (15.5) | 14,369 (17.8) | 573 (23.5) | 329 (18.8) |
| Max previous<br>viral test<br>results<br>(positive ><br>negative ><br>unknown) (%) | Not tested | 10.2 | 80,834 (84.3) | 7,916 (71.4) | 69,683 (86.4) | 1,890 (77.5) | 1,345 (76.8) |
|  | Non-definitive<br>result | 14.4 | 138 (0.1) | 19 (0.2) | 113 (0.1) | 2 (0.1) | 4 (0.2) |
|  | Negative | 8.4 | 12,313 (12.8) | 971 (8.8) | 10,622 (13.2) | 405 (16.6) | 315 (18.0) |
|  | Positive | 90.5 | 2,645 (2.8) | 2,188 (19.7) | 229 (0.3) | 141 (5.8) | 87 (5.0) |

BMI=body mass index; IQR=interquartile range.
<sup>a</sup> Indexed on procedural code without corresponding laboratory result
<sup>b</sup> 0, 25th, 50th, 75th, and 100th percentiles.
<sup>c</sup> Patients with ≤365 days of database history were not considered for comorbidities.

Among C3, the overall PR was 12%. Univariate results suggest that the highest PRs occurred among patients from the Northeast (18%), tested in the hospital (21%), and/or had Medicaid insurance (25%).

Among patients who received a definitive antibody test, 17% had a CLI recorded during the prior 6 weeks, of which, 58% occurred in the 7 days pre-index (Table 3).

Approximately 16% of these patients had received a prior viral test, of whom 18% were antibody positive (3% of C3).

### Concordance between Viral and Antibody Tests

Among patients who received a definitive viral test followed by (or on the same day as) a definitive antibody test (n=13,975), the overall concordance was 91% for both negative and positive results (Table 4). Concordance ranged from 89%-92% for negative and 73%-95% for positive results, respectively, according to the number of weeks between the tests. Discordant test results occurred in 9% of patients with little variability over time for patients with an initial negative viral test; however, discordant results were more common among patients who had an initial viral positive result and received an antibody test within 2 weeks (27.3%, 26.7%, and 15.1%; 0-, 1-, and 2-week intervals, respectively).

**Table 4.**
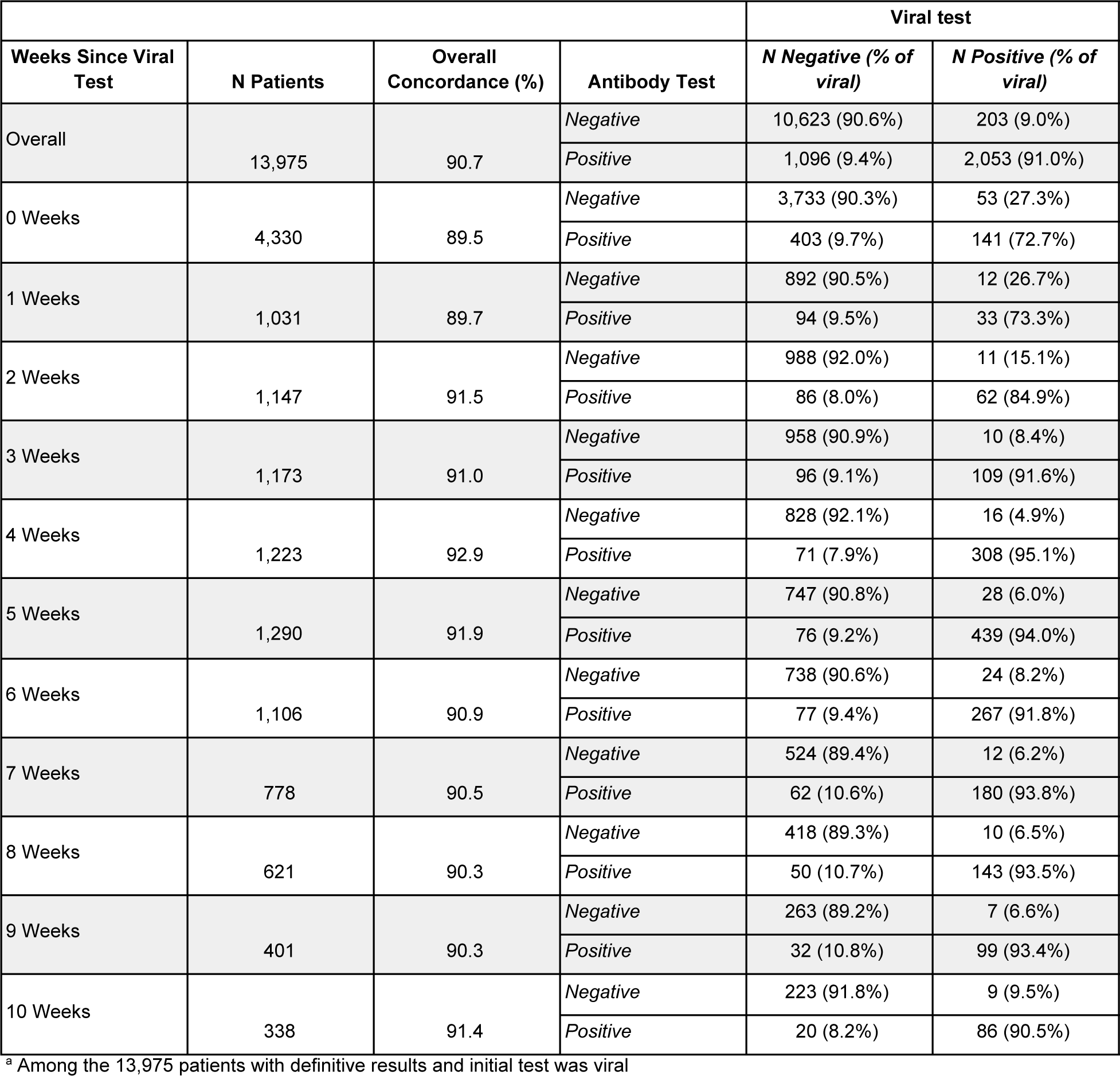
Concordance between SARS-CoV-2 Viral and Antibody Tests^a^

| Weeks Since Viral Test | N Patients | Overall Concordance (%) | Antibody Test | Viral test |  |
| --- | --- | --- | --- | --- | --- |
|  |  |  |  | N Negative (% of viral) | N Positive (% of viral) |
| Overall | 13,975 | 90.7 | Negative | 10,623 (90.6%) | 203 (9.0%) |
|  |  |  | Positive | 1,096 (9.4%) | 2,053 (91.0%) |
| 0 Weeks | 4,330 | 89.5 | Negative | 3,733 (90.3%) | 53 (27.3%) |
|  |  |  | Positive | 403 (9.7%) | 141 (72.7%) |
| 1 Weeks | 1,031 | 89.7 | Negative | 892 (90.5%) | 12 (26.7%) |
|  |  |  | Positive | 94 (9.5%) | 33 (73.3%) |
| 2 Weeks | 1,147 | 91.5 | Negative | 988 (92.0%) | 11 (15.1%) |
|  |  |  | Positive | 86 (8.0%) | 62 (84.9%) |
| 3 Weeks | 1,173 | 91.0 | Negative | 958 (90.9%) | 10 (8.4%) |
|  |  |  | Positive | 96 (9.1%) | 109 (91.6%) |
| 4 Weeks | 1,223 | 92.9 | Negative | 828 (92.1%) | 16 (4.9%) |
|  |  |  | Positive | 71 (7.9%) | 308 (95.1%) |
| 5 Weeks | 1,290 | 91.9 | Negative | 747 (90.8%) | 28 (6.0%) |
|  |  |  | Positive | 76 (9.2%) | 439 (94.0%) |
| 6 Weeks | 1,106 | 90.9 | Negative | 738 (90.6%) | 24 (8.2%) |
|  |  |  | Positive | 77 (9.4%) | 267 (91.8%) |
| 7 Weeks | 778 | 90.5 | Negative | 524 (89.4%) | 12 (6.2%) |
|  |  |  | Positive | 62 (10.6%) | 180 (93.8%) |
| 8 Weeks | 621 | 90.3 | Negative | 418 (89.3%) | 10 (6.5%) |
|  |  |  | Positive | 50 (10.7%) | 143 (93.5%) |
| 9 Weeks | 401 | 90.3 | Negative | 263 (89.2%) | 7 (6.6%) |
|  |  |  | Positive | 32 (10.8%) | 99 (93.4%) |
| 10 Weeks | 338 | 91.4 | Negative | 223 (91.8%) | 9 (9.5%) |
|  |  |  | Positive | 20 (8.2%) | 86 (90.5%) |

<sup>a</sup> Among the 13,975 patients with definitive results and initial test was viral

### Pediatric Patients (<18 Years)

Pediatric patients were underrepresented in our study population (C1=4.8%, C2=4.9%, C3=3.1%) relative to both the underlying Optum (15.4%) and census populations (22.3%). Pediatric patients had a viral PR of 6% and, while the PR was consistently lower among pediatric patients compared to the overall C2 population (9%), Hispanic, underweight, and Medicaid insured children were more likely to be tested (Table 2; Supplement I). Pediatric patients had an antibody PR of 17% compared to overall C3 PR of 12% with a particularly high PR among patients with Medicaid insurance, prior positive viral test, and >7 days since CLI (Supplement J). Of note, those underweight and/or hospitalized were more frequently tested compared to the overall C3 (Table 3; Supplement J). Among those who received both a viral and antibody test (n=405), concordance was 85% for negative results and 80% for positive results (Supplement K).

### Multivariable Logistic Model Results

#### Cohort 2

Of the 806,510 patients in C2, 657,112 (81%) fit the criteria for the model of viral test positivity.

#### Sociodemographic

The odds of a positive test were not meaningfully different among adults of different ages, when mutually adjusted for other factors (Figure 5, Supplement L); however, pediatric patients had a markedly decreased odds of a positive test relative to young adults (18-34 years) (odds ratio (OR)=0.60, 95% confidence interval (CI) 0.57, 0.64). Consistently decreased odds were observed among pediatric patients in an assessment of an interaction between age and number of symptoms (data not shown).

**Figure 5.**
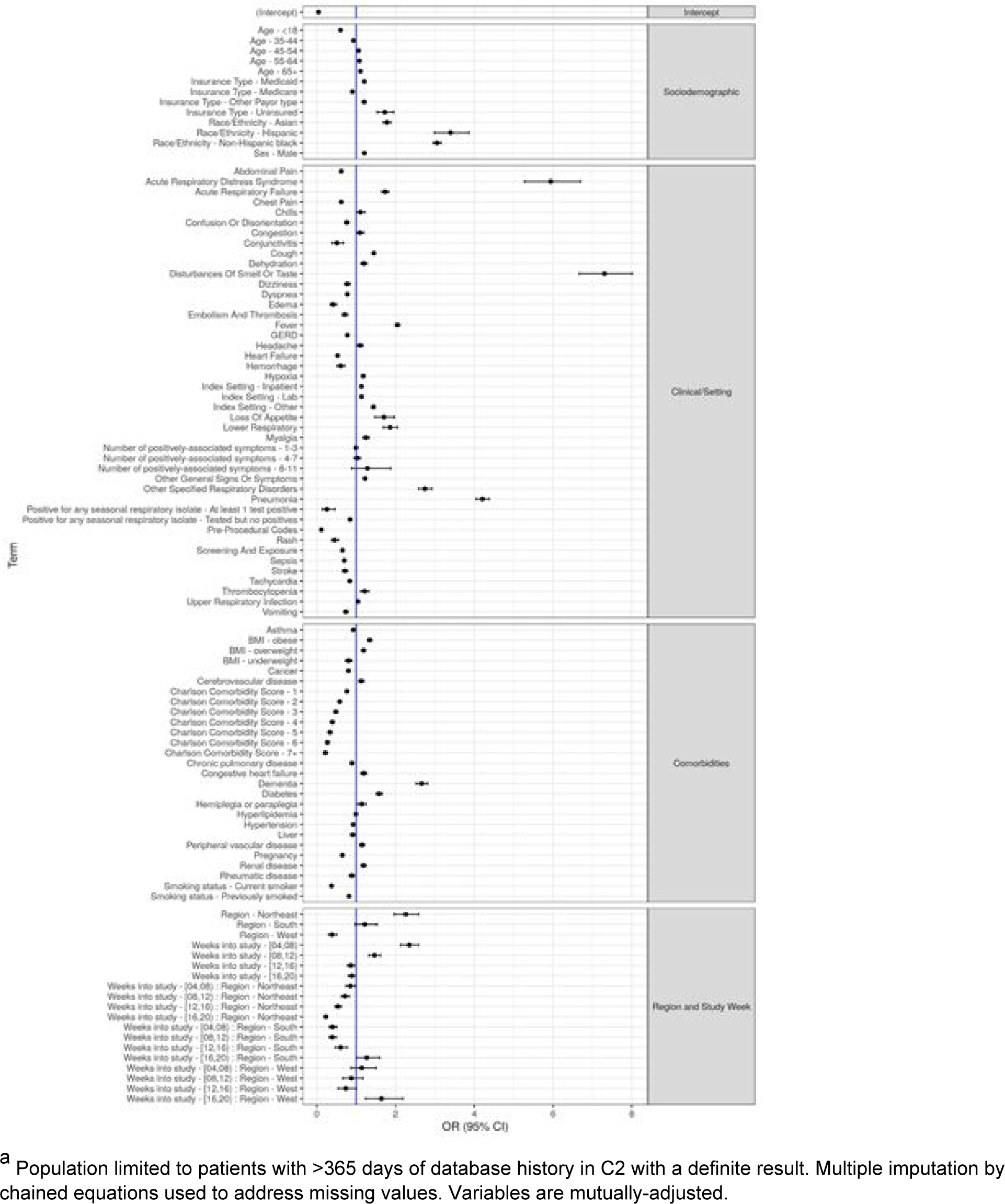
Odds Ratios and 95% Confidence Intervals of SARS-CoV-2 Positive Viral Test^a^

Hispanic, non-Hispanic Black, and Asian patients had increased odds of viral positivity compared to non-Hispanic Whites (OR and 95% CI=3.40, 2.99, 3.86; 3.06, 2.96, 3.16; and 1.78, 1.69, 1.89, respectively). Uninsured patients and those with Medicaid had moderately increased odds of testing positive, while Medicare patients had a decreased odds, relative to patients with commercial insurance (OR and 95% CI=1.73, 1.53, 1.95; 1.21, 1.17, 1.25; and 0.91, 0.87, 0.94, respectively). Males had an increased odds of positivity relative to females (OR=1.21, 95% CI 1.19, 1.24).

#### Clinical/Setting

To assess severity, symptomatic status, and viral test positivity, we investigated the importance of healthcare setting and symptoms. We observed modest significant associations between healthcare setting and positivity (OR=1.14, 95% CI 1.10, 1.17) for patients tested in the hospital compared to outpatient setting.

We identified several symptoms/co-occurring diagnoses that were significantly associated with a positive test, including: disturbance of smell and/or taste (OR=7.31, 95% CI 6.67, 8.01), acute respiratory distress (OR=5.94, 95% CI 5.27, 6.70), pneumonia (OR=4.21, 95% CI 4.04, 4.38), lower respiratory infection (OR=1.86, 95% CI 1.69, 2.05), loss of appetite (OR=1.71 95% CI 1.48, 1.97), and cough (OR=1.45, 95% CI 1.41, 1.49). Other conditions with modestly significant positive and negative associations are shown in Figure 5 and Supplement L. Patients who had a diagnostic code reported for (pre)procedural test or screening were less likely to test positive than those without such a code (OR and 95% CI=0.12, 0.11, 0.13; 0.66, 0.64, 0.67, respectively). Further, patients tested for a non-SARS-CoV-2 respiratory isolate were less likely to test positive for SARS-CoV-2 compared to patients who were not tested (OR and 95% CI=0.85, 0.82, 0.87 tested but negative; 0.26, 0.14, 0.46 tested and positive for ≥1 pathogen).

### Comorbidities

We found an inverse dose response relationship between CCI score and viral positivity indicating that patients with more comorbidities had lower odds of a positive result.

However, assessment of individual conditions identified an increased odds of a positive test among patients with dementia (OR=2.66, 95% CI 2.52, 2.82) and diabetes (OR=1.59, 95% CI 1.51, 1.67) that persisted in mutually exclusive exploratory models restricted to 65+ suggesting limited residual confounding by age (data not shown). It is possible that the increased odds of a positive test among dementia patients may be related to increased exposure due to institutional living and/or caregivers but we were unable to directly assess this within our data. Very modest significant associations were identified for patients with conditions such as renal disease (OR=1.19, 95% CI 1.13, 1.25) and congestive heart failure (OR=1.20, 95% CI 1.12, 1.27) (Figure 5; Supplement L). Further, overweight and obese patients had an increased odds of a positive test (OR=1.19, 95% CI 1.14, 1.24 and OR=1.34, 95% CI 1.29, 1.40, respectively) relative to patients with normal BMI. We identified several comorbidities with negative associations, such as cancer, asthma and rheumatic disease; however, effect estimates were generally modest.

### Region and Study Week

There was a significant interaction between region and study week on the odds of a positive test. Our model reflects high transmission in the Northeast region early in the pandemic with attenuated effects in this region over time suggesting transmission control relative to other regions such as the Midwest (Figure 5). Conversely, the South experienced increased ORs in interaction terms with study week, while the West experienced a J-shaped curve with the highest OR in the final study week.

### Cohort 3

Of the 95,930 patients in C3, 83,184 (87%) fit the eligibility criteria for the logistic model of antibody test positivity (Figure 6; Supplement M).

**Figure 6.**
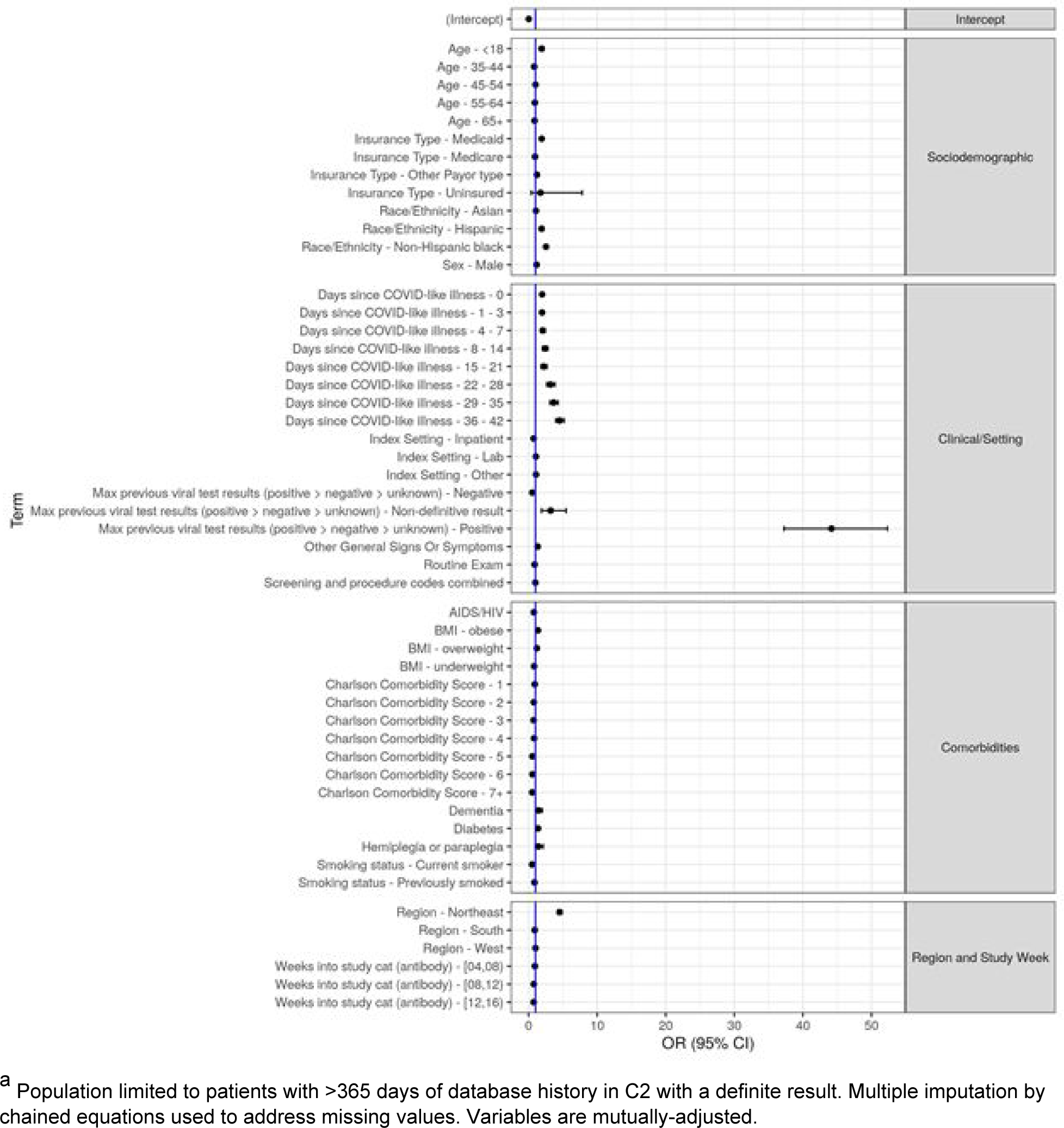
Odds Ratios and 95% Confidence Intervals of SARS-CoV-2 Positive Antibody Test ^a^

### Sociodemographic

We detected increased odds of a positive antibody test among patients who: were <18 years (OR=1.90, 95% CI 1.62, 2.23 compared to 18-34 years); had Medicaid or no insurance (OR=1.91, 95% CI 1.75, 2.08 and 1.73, 95% CI 0.38, 7.82 compared to patients with commercial insurance); were non-Hispanic Black or Hispanic patients (OR=2.54, 95% CI 2.32, 2.77 and OR=1.90, 95% CI 1.63, 2.21 compared to non-Hispanic Whites); lived in the Northeast region (OR=4.53, 95% CI 4.22, 4.86 compared to the Midwest); or were male (OR=1.16, 95% CI 1.11, 1.22).

### Clinical/Setting

As expected, the odds of a positive antibody test among patients with a prior viral positive test was particularly high (OR=44.16, 95% CI 37.26, 52.33), and low (OR=0.54, 95% CI 0.49, 0.58) for patients with a prior negative viral test compared to patients who did not receive a prior viral test. The odds of a positive antibody test increased with the number of days since CLI (ORs ranged from 1.97, 95% CI 1.81, 2.14 to 4.53, 95% CI 3.97, 5.18) relative to patients without a prior CLI.

The odds of a positive test were lower among patients tested in the hospital relative to ambulatory setting (OR=0.73, 95% CI 0.57, 0.92). Similar to C2 findings, C3 patients who had a routine general medical exam recorded were less likely to have a positive test result compared to patients without such a code (OR=0.88, 95% CI 0.82, 0.94).

### Comorbidities

Our assessment of the relationship between CCI score and antibody test positivity was consistent with results from C2. Specifically, we identified an inverse dose effect between CCI score and test positivity (OR ranged from 0.90, 95% CI 0.84, 0.95 to 0.52, 95% CI 0.34, 0.80 as the number of comorbidities increased from 1 to ˃7 relative to patients with a CCI score of 0). The significantly positive association between dementia, diabetes, and BMI persisted in C3.

## CONCLUSION

We characterized sociodemographic and clinical factors associated with SARS-CoV-2 testing and positivity in over 800,000 patients across the US. Our study provides real-world clinical data that complements surveillance data and identifies novel findings related to both patients and testing.

Our findings support the need for greater viral testing in minority populations due to low proportion of testing and high PRs (Table 2; Figure 2), as well as heightened positivity in our multivariable model (Figure 5). However, Hispanic patients were underrepresented in the Optum population relative to the census population (Table 1). Our findings related to minority populations and sub-optimal health insurance are consistent with prior research demonstrating the importance of societal and economic inequities, differential exposure, access to tests over the course of the pandemic [12, 13, 16, 20, 21, 22, 23, 24, 25, 26, 27, 28, 29].

Our assessment of both viral and antibody tests provides novel data regarding real-world evidence of test concordance among patients who received both tests. While concordance was high, the time interval between tests should be considered as viral and antibody tests taken <2 weeks apart had lower concordance. This may reflect seroconversion time between symptom onset detectable antibodies [30]. High concordance between tests is notable given variable test characteristics and our inability to directly assess sensitivity and specificity within, and between, tests within available EHRs.

We identified notable differences in PRs among pediatric patients depending upon the type of test compared to the overall C2 and C3 patients. Concordance between viral and antibody tests among pediatric patients was slightly lower than for all patients. Our findings may reflect variable testing patterns and results due to differences in clinical presentation, symptomatic status, severity, immune response, and detection in children which is consistent with national surveillance data (Figure 7) [31, 32, 33, 34].

**Figure 7.**
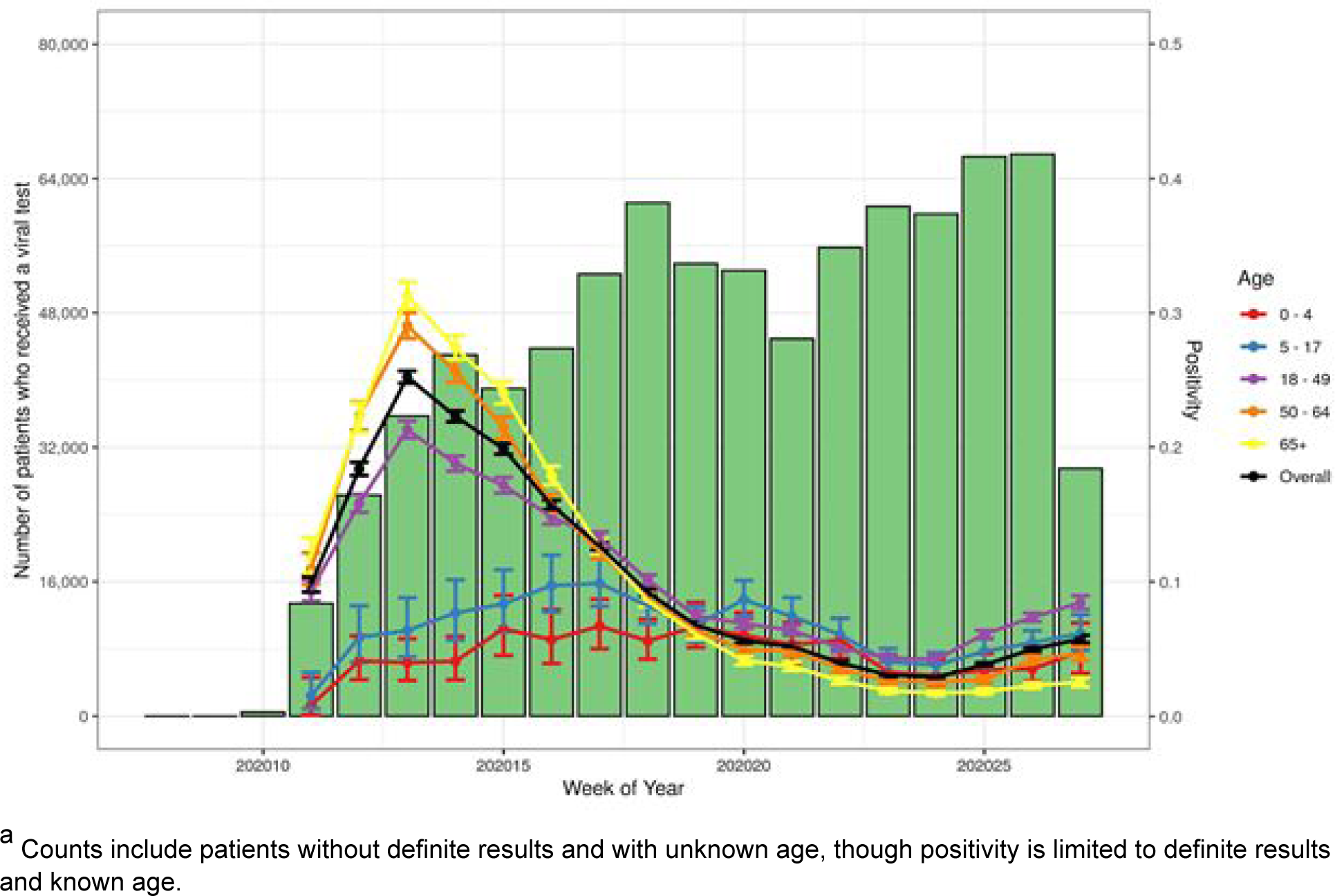
Number of Patients Tested and Percent Positive for Initial SARS-CoV-2 Viral Test by Age^a^

While EHRs may lack sensitivity for capturing symptoms, our findings support the importance of symptoms such as olfactory dysfunction, fever, cough and lower respiratory infections, and viral positivity [14, 35, 36]. However, we also report negative associations between co-occurring symptoms and positivity that may reflect true relationships, conservative testing practices among vulnerable patients, or ambiguous symptoms that lead to testing but are not COVID-19-related or present during periods of suboptimal test performance.

While patients with poor underlying health may be at higher-risk of severe COVID-19, the negative association between multi-morbidities and test positivity is consistent with prior studies and may reflect reduced exposure due to behavioral changes and/or lower testing thresholds [15, 37, 38, 39, 40]. Exceptions were noted for patients with obesity, diabetes, and dementia, which are also associated with severe disease [41, 42, 43].

We recognize that differential access to, and thresholds for, testing exist and that not all EHR records are complete [44]. We did not require a minimum follow-up period for inclusion into our study given our interest in testing results that did not require extended follow-up and our intent was to be as inclusive as possible. We assessed the representativeness of our study population to the Optum and census populations, and utilized days since CLI as a proxy for time since symptom onset. While we compared our study population to source populations, we were unable to directly describe the base population due to the nature of the de-identified Optum COVID-19 EHR data, Nor were we able to assess test characteristics according to timing of infection or symptom onset, test manufacturer, or specimen type [8, 39].

In summary, this large national study systematically identified sociodemographic and clinical factors associated with SARS-CoV-2 testing and positivity. Our findings identify the need for additional testing among minority patients and provide novel findings related to antibody testing.

## List of Abbreviations

BMI: Body Mass Index
C1: Cohort 1
C2: Cohort 2
C3: Cohort 3
CCI: Charlson Comorbidity Index
CDC: Centers for Disease Control and Prevention
CI: Confidence Interval
CLI: COVID-like Illness
COVID-19: Coronavirus Disease 2019
CPT: Current Procedural Terminology
EHR: Electronic Health Record
HCPC: Healthcare Common Procedure Coding System
HIPAA: Health Insurance Portability and Accountability Act LOINC Logical Observation Identifiers Names and Codes
OR: Odds Ratio
PLA: Proprietary Laboratory Analyses
PR: Positivity Rate
SARS-CoV-2: Severe Acute Respiratory Syndrome Coronavirus 2 US United States

## Supporting information

Supplementary Material COVID-19 Testing and Positivity

## Data Availability

The data that support the findings of this study are available from Optum but restrictions apply to the availability of these data, which were used under license for the current study, and so are not publicly available. Data are however available from the authors upon reasonable request and with permission of Optum.

## Acknowledgements

The authors thank Xin Chen, Devika Chawla, Vince Yau, Tripthi Kamath, and Jenny Chia for their review and input into this study.

