## Supplementary Material COVID-19 Testing and Positivity for "Viral And Antibody Testing For Coronavirus Disease 2019 (Covid-19): Factors Associated With Positivity In Electronic Health Records From The United States"

Lisa Lindsay, Personalized Healthcare, Data Science, Genentech Inc., South San Francisco, USA
Matthew H. Secrest, Personalized Healthcare, Data Science, Genentech Inc., South San Francisco, USA
Shemra Rizzo, Personalized Healthcare, Data Science, Genentech Inc., South San Francisco, USA
Dan Keebler, Personalized Healthcare, Data Science, Genentech Inc., South San Francisco, USA
Fei Yang, Data Services, Roche Information Solutions, Roche Diagnostics, Basel, Switzerland
Larry W. Tsai, Product Development, Genentech Inc., South San Francisco, USA

**Keywords.** COVID-19; SARS-CoV-2; testing; positivity; viral; antibody; concordance; temporality; EHR

**Running title:** COVID-19 Testing and Positivity

**Contact information for corresponding author**

Lisa Lindsay, PhD, MPH

Genentech, Inc., 1 DNA Way, South San Francisco, California 94080, USA

Supplement A. Study Cohort Definition

| Codes Used to Identify Patient Cohorts | | |
|---|---|---|
| Laboratory and Procedural Codes | | |
| Cohort 1: SARS-CoV-2 Lab Tested | Cohort 2: SARS-CoV-2 Viral Tested | Cohort 3: SARS-CoV-2 Antibody Tested |
| 94534-5, 94500-6, 94503-0,<br><br>94505-5, 94506-3, 94504-8,<br><br>94507-1, 94509-7, 94508-9,<br><br>94510-5, 94308-4, 94309-2,<br><br>94311-8, 94312-6, 94308-4,<br><br>94309-2, 94311-8, 94312-6,<br><br>94314-2, 94315-9, 94316-7,<br><br>94533-7, 94531-1, 94547-7,<br><br>94564-2, 94562-6, 94563-4,<br><br>94564-2, 94562-6, 94563-4,<br><br>94565-9, 94307-6, 94559-2,<br><br>94306-8, 94558-4, 94511-3<br><br>86328, 86769, 87635, 86769,<br><br>G2023, G2024, U0001-4,<br><br>0224U, 0202U, 0223U | 94534-5, 94500-6, 94509-7,<br><br>94510-5, 94308-4, 4309-2,<br><br>94311-8, 4312-6, 94314-2,<br><br>4315-9, 94316-7, 4533-7,<br><br>94531-1, 4565-9, 94307-6,<br><br>4559-2, 94306-8, 4558-4,<br><br>94511-3, 87635, 0001, U0002,<br><br>U0003, U0004, 0202U, 0223U<br><br>If a test occurs on a date or encounter with a viral HCPCS/CPT ('87635', 'U0001', 'U0002', 'U0003', 'U0004', ‘0202U’, ‘0223U’) and no antibody HCPCS/CPT ('86328', '86769', ‘0224U’), assign viral | 94503-0, 94505-5, 94506-3,<br><br>94504-8, 94507-1, 94508-9,<br><br>94547-7, 94564-2, 94562-6,<br><br>94563-4, 86328, 86769, 0224U<br><br>If a test occurs on a date or encounter with an antibody procedure code ('86328', '86769', ‘0224U’) and no viral HCPCS/CPT ('87635', 'U0001', 'U0002', 'U0003', 'U0004', ‘0202U’, ‘0223U’), assign antibody |
| String Search | | |
| Identify labs via regular expressions applied to test names<br><br>*coronavirus* and *2, 19*, or *novel* but no terms to identify other coronaviruses (*human/nl63/hku1/oc43/229e/not/non*)<br><br>*covid* and no other irrelevant terms (*not/non/symptomatic/eval*)<br><br>*sars* and *2*<br><br>Exact match of ‘*sars cov pcr’* | If a test has a name consistent with a viral test and inconsistent with an antibody test, or if a test indicates a specimen type that is consistent with viral test (eg ‘naso’, ‘oro’, ‘swab’, ‘trachea’, ‘saliva’), assign viral | If a test has a name consistent with antibody and inconsistent with viral (eg ‘ab’, ‘anti’, ‘igg’, ‘igm’, without ‘pcr’), assign antibody |

| |
|---|
| Exclude irrelevant tests based on test results<br><br>*Pregnancy, spirochetes, ebv nuclear antigen, hbv, cerebrospinal, cervix, influenza, malaria, anemia* |

Supplement B. Description and Codes used to Identify Symptoms/Co-Occurring Diagnoses

| Symptom | Definition |
|---|---|
| **Vitals** | |
| Abnormal EKG | ICD10: R9431<br>ICD9: 79431 |
| Tachycardia | Any PULSE >100 bpm or ICD10 code: R000 or ICD9 code: 7850 |
| High blood pressure | High blood pressure is any systolic > 190 or diastolic > 90 or ICD10: I10 or ICD9: 4010, 4011, 4019 |
| Hypoxia/<br>Oxygen Saturation / Cyanosis | ICD10 codes: ('R0902', '79902', 'J9691', 'J9611', 'J9621', 'I2723', ‘R230’),<br><br>ICD9 codes: (‘79902’, ‘7825’)<br><br>Any pulse oximetry < 93 |
| **Systemic** | |
| Malaise | ICD10: R5381<br>ICD9: 78079 |
| Fever | Any TEMP observation >38°C or ICD10 code R509 or ICD9 code 78060 |
| Rash | ICD10: R21<br>ICD9: 7821 |
| Fatigue | ICD10: R5383<br>ICD9: 78079 |
| Myalgia | ICD10: M791x<br>ICD9: 7291 |
| Edema/swelling | ICD10: R609<br>ICD9: 7823 |
| Chilblains/red patches | ICD10: T691x<br>ICD9: 9915 |
| Sepsis | Codes for sepsis:<br>ICD10: A4189-A419, R652x<br>ICD9: 99591-99592 |
| Chills | ICD10: R05<br>ICD9: 7862 |

| | |
|---|---|
| **Respiratory** | |
| Congestion | ICD10: R0981<br>ICD9: 47819 |
| Shortness of breath / Dyspnea | ICD10 code: R060x<br>ICD9 code: 78609, 78602, 78605, 51882 |
| Upper respiratory infection | ICD10 code: J00-J01, J028-9, J038-9, J04-J05<br>ICD9 code: 460-465 |
| Acute respiratory failure | ICD10 code: J960x, J962x, J969x<br>ICD9 code: 51881, 51884 |
| Cough | ICD10 code: R05<br>ICD9 code: 7862 |
| Bronchitis | ICD10 code: J208-9, J40<br>ICD9 code: 4660, 490 |
| Bronchiolitis | ICD10 code: J218-9<br>ICD9 code: 46619 |
| Pneumonia | ICD10 codes: J12-18<br>ICD9 codes:481-6 |
| Lower respiratory infection | ICD10 codes: J22 |
| Acute respiratory distress syndrome | ICD10 codes: J80<br>ICD9 code: 51882 |
| Other specified respiratory disorders | ICD10 codes: J988<br>ICD9 codes: 5198 |
| **Cardiovascular/ Hematologic** | |
| Syncope/fainting | ICD10: R55<br>ICD9: 7802 |
| Chest pain | ICD10 code: R079<br>ICD9 code: 78650 |
| Hyperlipidemia | ICD10 code: E785<br>ICD9 code: 2724 |
| Hypertension | ICD10: I10<br>ICD9: 4010, 4011, 4019 |
| Respiratory and Unspecified Bleeding | ICD10: R04x, R58<br>ICD9: 4590, 784x, 786x |
| Disseminated intravascular coagulation | ICD10: D65<br>ICD9: 2866 |
| Thrombocytopenia | ICD10: D696<br>ICD9: 2875 |

| | |
|---|---|
| Embolism and thrombosis | ICD10: I74-I76, I82, I26,<br>ICD9: 444-5, 449, 453, 415 |
| Stroke | ICD10: I60-I69<br>ICD9: 430-438 |
| Acute heart failure | ICD10: I50x<br>ICD9: 428x |
| **Gastrointestinal** | |
| Diarrhea | ICD10: R197<br>ICD9: 78791 |
| Nausea | ICD10: R110, R112<br>ICD9: 78701-78702 |
| Vomiting | ICD10: R111-R112<br>ICD9: 78701, 78703 |
| Abdominal pain | ICD10: R10x<br>ICD9: 7890x, 7896x |
| Dehydration | ICD10: E860<br>ICD9: 27651 |
| Loss of appetite | ICD10: R630<br>ICD9: 7830 |
| GERD | ICD10: K21x<br>ICD9: 53081 |
| **Renal** | |
| Acute kidney failure | ICD10: N17x, N19x<br>ICD9: 584x, 586x |
| **Neurologic** | |
| Headache | ICD10: R51<br>ICD9: 7840 |
| Confusion/ disorientation | ICD10: R410, R4182<br>ICD9: 78097 |
| Disturbance of smell/taste (anosmia, parageusia, parosmia) | ICD10: R43x<br>ICD9: 7811 |
| Hypersomnia | ICD10: G4710-3, G4719<br>ICD9: 78053-4, 32710-3, 32719 |
| Dizziness | ICD10: R42<br>ICD9: 7804 |
| **Ophthalmologic/Eye Involvement** | |

| | |
|---|---|
| Conjunctivitis | ICD10: H10x<br>ICD9: 3720-3723x |
| Light sensitivity / visual discomfort | ICD10: H53149<br>ICD9: 36813 |
| Edema of the eye | ICD10: H0284x<br>ICD9: 37482 |
| **General** | |
| Other general signs/symptoms [new] | ICD10: R6889<br>ICD9: 78099 |
| COVID-related U codes | ICD10: U071-2 |
| COVID-related B code | B9729 |
| **Pre-procedural** | |
| Pre-procedural / pre-operative codes [new] | ICD10: Z0181x<br>ICD9: V7281-4 |
| **Screening / exposure codes** | |
| Screening / exposure codes [new] | ICD10: Z1159, Z20828, Z03818<br>ICD9: V7389, V7399, V0179, V7183 |

Supplement C. COVID-like Illness Definition

The definition of CLI will be considered if the following criteria are met on the same day:

- 2+ of the following symptoms
    - Fever with or without rigors (based on temperature or ICD codes R509 or 78060)
    - Chills (ICD codes R6883 or 78064)
    - Myalgia (ICD codes M791x or 7291)
    - Headache (ICD codes R51 or 7840)
    - Sore Throat (ICD codes J028-J029 or 462)
    - Nausea or Vomiting (ICD codes R11x or 78701-3)
    - Diarrhea (ICD codes R197 or 78791)
    - Fatigue (ICD codes R5383 or 78079)
    - Congestion or Runny Nose (ICD codes R0981 or 47819)

OR

- 1+ of the following symptoms
    - Cough (ICD codes R05 or 7862)
    - Shortness of Breath / difficulty breathing (ICD codes R06x, 78609, 78602, 78605, 51882)
    - New Olfactory or Taste Disorder (R43x or 7811) (among those without such diagnosis in prior 43-180 days)

OR

- Pneumonia (J12x-8x, 481-6)
- Acute Respiratory Distress Syndrome (J80, 51882)
- COVID-19 virus identified, diagnosis confirmed by lab testing (U072)
- COVID-19 not identified, diagnosis but lab inconclusive or not available (U071)
- Other coronavirus as the cause of diseases classified elsewhere (B9729)
- Acute bronchitis due to other specified organisms (J208-9, 4660)
- Bronchitis, not specified as acute or chronic (J40, 490)
- Unspecified acute lower respiratory infection (J22)
- Other specified respiratory disorders (J988, 5198)

*This definitive was adapted from the* Clinical Criteria Endorsed by CDC & Council of State and Territorial Epidemiologists August 2020 *(see: https://wwwn.cdc.gov/nndss/conditions/coronavirus-disease-2019-covid-19/case-definition/2020/08/05/), with the addition of symptom codes (J codes) used to create the Optum® COVID dataset and the COVID diagnostic codes (U071, U072, and B9729)*

Supplement D. Description and Codes used to Identify Comorbidities / Existing Conditions[a]

| **Comorbidity / Existing Condition** | **Definition** |
|---|---|
| BMI | Closest valid (between 5 and 60 kg/m^2) BMI recording within 365 days prior to the index date.<br><br>Underweight: < 18.5 kg/m^2<br>Normal: [18.5, 24.0) kg/m^2<br>Overweight: [24.0, 30) kg/m^2<br>Obese: >30 kg/m^2 |
| Smoking status | Closest recording within 365 days prior to index. |
| Charlson Comorbidity Score [a] | |
| Myocardial Infarction | ICD codes: I21x, I22x, I252, 410x, 412x |
| Congestive heart failure | ICD codes: I099, I110, I130, I132, I255, I420, I425-I429, I43x, I50x, P290, 39891, 40201, 40211, 40291, 40401, 40403, 40411, 40413, 40491, 40493, 4254-4259, 428x |
| Peripheral vascular disease | ICD codes: I70x, I71x, I731, I738, I739, I771, I790, I792, K551, K558, K559, Z958, Z959,0930, 4373, 440x, 441x, 4431-4439, 4471, 5571, 5579, V434 |
| Cerebrovascular disease | ICD codes: G45x, G46x, H340, I60x-I69x, 36234, 430x-438x |
| Dementia | ICD codes: F00x-F03x, F051, G30x, G311, 290x, 2941, 3312 |
| Chronic pulmonary disease | ICD codes: I278, I279, J40x-J47x, J60x-J67x, J684, J701, J703, 4168, 4169, 490x-505x, 5064, 5081, 5088 |
| Rheumatic disease | ICD codes: M05x, M06x, M315, M32x-M34x, M351, M353, M360, 4465, 7100-7104, 7140-7142, 7148, 725x |
| Peptic ulcer disease | ICD codes: K25x-K28x, 531x-534x |
| Mild liver disease | ICD codes: B18x, K700-K703, K709, K713-K715, K717, K73x, K74x, K760, K762-K764, K768, K769, Z944, 07022, 07023, 07032, 07033, 07044, 07054, 0706, 0709, 570x, 571x, 5733, 5734, 5738, 5739, V427 |
| Diabetes without chronic complications | ICD codes: E100, E10l, E106, E108, E109, E110, E111, E116, E118, E119, E120, E121, E126, |

| | |
|---|---|
| | E128, E129, E130, E131, E136, E138, E139, E140, E141, E146, E148, E149, 2500-2503, 2508, 2509 |
| Diabetes with chronic complications | ICD codes: E102-E105, E107, E112-E115, E117, E122-E125, E127, E132-E135, E137, E142-E145, E147, 2504-2507 |
| Hemiplegia or paraplegia | ICD codes: G041, G114, G801, G802, G81x, G82x, G830-G834, G839, 3341, 342x, 343x, 3440-3446, 3449 |
| Renal disease | ICD codes: I120, I131, N032-N037, N052-N057, N18x, N19x, N250, Z490-Z492, Z940, Z992, 40301, 40311, 40391, 40402, 40403, 40412, 40413, 40492, 40493, 582x, 5830-5837, 585x, 586x, 5880, V420, V451, V56x |
| Any malignancy except malignant neoplasm of skin | ICD codes: C00x-C26x, C30x-C34x, C37x-C41x, C43x, C45x-C58x, C60x-C76x, C81x-C85x, C88x, C90x-C97x, 140x-172x, 174x-1958, 200x-208x, 2386 |
| Moderate or severe liver disease | ICD codes: I850, I859, I864, I982, K704, K711, K721, K729, K765, K766, K767, 4560-4562, 5722-5728 |
| Metastatic solid tumor | ICD codes: C77x-C80x, 196x-199x |
| AIDS/HIV | ICD codes: B20x-B22x, B24x, 042x-044x |
| Combined Charlson Comorbidity Index measures [a] | |
| Any liver disease | Mild liver disease or moderate liver disease (above) |
| Any cancer | Any malignancy or any metastatic solid tumor (above) |
| Any diabetes | Diabetes with complications or diabetes without complications (above) |
| Other Comorbidities / Co-occurring conditions | |
| Pregnancy | Evaluated within the 0-180 days prior to the index date among patients with >180 days of database history<br><br>ICD codes: O09x, Z34x-Z36x, V22x-V23x, V28x, 659x |

| | |
|---|---|
| Asthma[b] | ICD codes: J45x-J46, 493x |
| Hypertension [b] | ICD codes: I10-I15x, 401x-405x |
| Hyperlipidemia [b] | ICD codes: E78x, 272x |

[a] ≥1 diagnosis code was sufficient to consider the patient as having the condition
[b] Evaluated within the 0-365 days prior to the index date among patients with >365 days of database history.

Supplement E. Missingness Proportion by Variables Included in Multivariable Model for SARS-CoV-2 Positivity via Viral Test (Cohort 2)

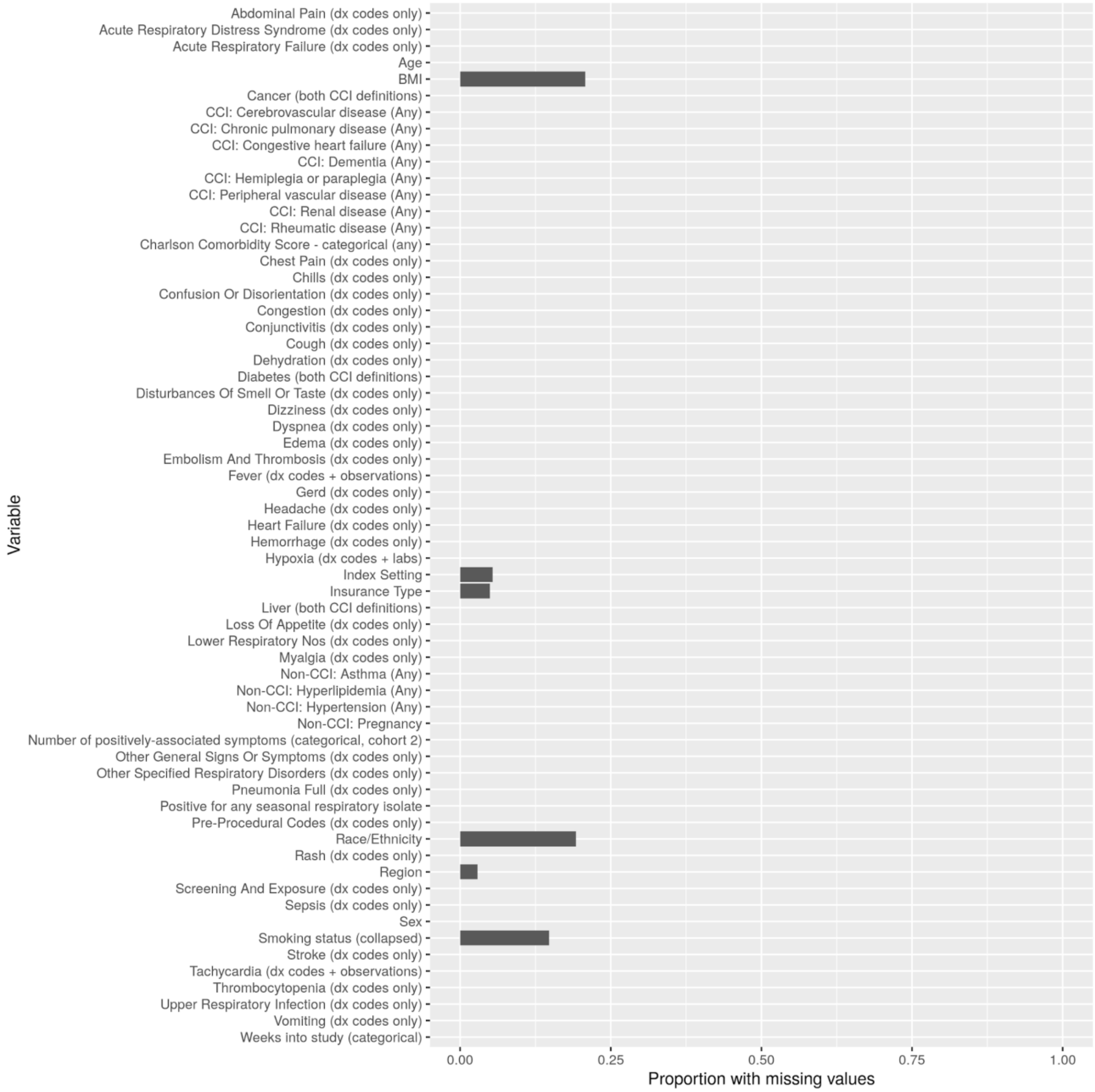

Supplement F. Missingness Proportion by Variables Included in Multivariable Model for SARS-CoV-2 Positivity via Antibody Test (Cohort 3)

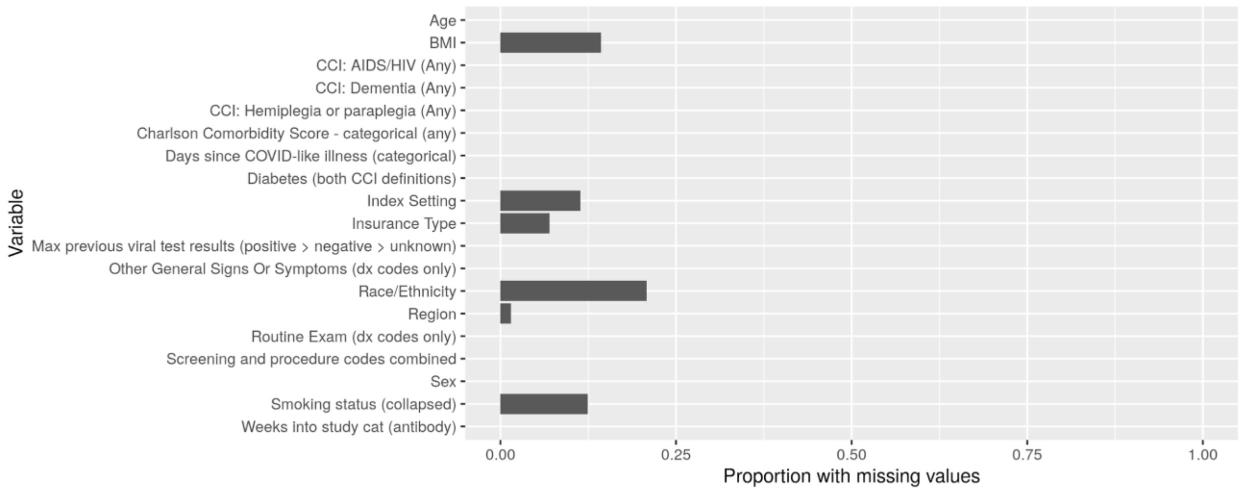

Supplement G. Distribution of Symptoms by Cohort

| Variable | Level | Cohort 1 | Cohort 2 | Cohort 3 |
|---|---|---|---|---|
| n | | 891,754 | 806,510 | 95,930 |
| Days since COVID-like illness (%) | No COVID-like illness observed | 688,288 (77.2) | 614,872 (76.2) | 79,264 (82.6) |
| | 0 | 130,832 (14.7) | 125,608 (15.6) | 5,879 (6.1) |
| | 1 - 3 | 27,520 (3.1) | 25,845 (3.2) | 2,307 (2.4) |
| | 4 - 7 | 8,286 (0.9) | 7,319 (0.9) | 1,373 (1.4) |
| | 8 - 14 | 8,044 (0.9) | 7,169 (0.9) | 1,347 (1.4) |
| | 15 - 21 | 6,413 (0.7) | 5,756 (0.7) | 1,059 (1.1) |
| | 22 - 28 | 6,447 (0.7) | 5,832 (0.7) | 1,052 (1.1) |
| | 29 - 35 | 6,829 (0.8) | 6,061 (0.8) | 1,489 (1.6) |
| | 36 - 42 | 9,095 (1.0) | 8,048 (1.0) | 2,160 (2.3) |
| Any vitals symptom (%) | FALSE | 466,194 (52.3) | 420,891 (52.2) | 51,538 (53.7) |
| | TRUE | 425,560 (47.7) | 385,619 (47.8) | 44,392 (46.3) |
| Any systemic symptom (%) | FALSE | 743,790 (83.4) | 663,688 (82.3) | 89,637 (93.4) |
| | TRUE | 147,964 (16.6) | 142,822 (17.7) | 6,293 (6.6) |
| Any cardiovascular symptom (%) | FALSE | 699,723 (78.5) | 632,930 (78.5) | 75,730 (78.9) |
| | TRUE | 192,031 (21.5) | 173,580 (21.5) | 20,200 (21.1) |
| Any gastrointestinal symptom (%) | FALSE | 777,275 (87.2) | 697,117 (86.4) | 90,208 (94.0) |
| | TRUE | 114,479 (12.8) | 109,393 (13.6) | 5,722 (6.0) |
| Any neurological symptom (%) | FALSE | 843,141 (94.5) | 759,494 (94.2) | 94,063 (98.1) |
| | TRUE | 48,613 (5.5) | 47,016 (5.8) | 1,867 (1.9) |
| Any ophthalmologic symptom (%) | FALSE | 890,251 (99.8) | 805,171 (99.8) | 95,720 (99.8) |
| | TRUE | 1,503 (0.2) | 1,339 (0.2) | 210 (0.2) |
| Any renal symptom (%) | FALSE | 857,104 (96.1) | 772,005 (95.7) | 95,577 (99.6) |
| | TRUE | 34,650 (3.9) | 34,505 (4.3) | 353 (0.4) |
| Any respiratory symptom (%) | FALSE | 632,908 (71.0) | 555,802 (68.9) | 85,849 (89.5) |
| | TRUE | 258,846 (29.0) | 250,708 (31.1) | 10,081 (10.5) |
| Any general sign or symptom (%) | FALSE | 813,100 (91.2) | 729,969 (90.5) | 92,839 (96.8) |
| | TRUE | 78,654 (8.8) | 76,541 (9.5) | 3,091 (3.2) |
| Hypertension (%) | FALSE | 484,837 (54.4) | 439,304 (54.5) | 51,762 (54.0) |
| | TRUE | 406,917 (45.6) | 367,206 (45.5) | 44,168 (46.0) |
| Fever (%) | FALSE | 793,555 (89.0) | 709,634 (88.0) | 94,048 (98.0) |
| | TRUE | 98,199 (11.0) | 96,876 (12.0) | 1,882 (2.0) |
| Tachycardia (%) | FALSE | 753,968 (84.5) | 671,011 (83.2) | 92,494 (96.4) |
| | TRUE | 137,786 (15.5) | 135,499 (16.8) | 3,436 (3.6) |
| Hypoxia (dx codes + labs) (%) | FALSE | 807,460 (90.5) | 722,894 (89.6) | 94,661 (98.7) |

| | | | | |
|---|---|---|---|---|
| | TRUE | 84,294 (9.5) | 83,616 (10.4) | 1,269 (1.3) |
| Abnormal EKG (%) | FALSE | 864,142 (96.9) | 779,552 (96.7) | 95,039 (99.1) |
| | TRUE | 27,612 (3.1) | 26,958 (3.3) | 891 (0.9) |
| Malaise (%) | FALSE | 877,327 (98.4) | 792,877 (98.3) | 95,004 (99.0) |
| | TRUE | 14,427 (1.6) | 13,633 (1.7) | 926 (1.0) |
| Chills (%) | FALSE | 886,255 (99.4) | 801,070 (99.3) | 95,854 (99.9) |
| | TRUE | 5,499 (0.6) | 5,440 (0.7) | 76 (0.1) |
| Rash (%) | FALSE | 888,257 (99.6) | 803,449 (99.6) | 95,306 (99.3) |
| | TRUE | 3,497 (0.4) | 3,061 (0.4) | 624 (0.7) |
| Fatigue (%) | FALSE | 865,122 (97.0) | 782,700 (97.0) | 92,711 (96.6) |
| | TRUE | 26,632 (3.0) | 23,810 (3.0) | 3,219 (3.4) |
| Myalgia (%) | FALSE | 878,703 (98.5) | 793,899 (98.4) | 95,430 (99.5) |
| | TRUE | 13,051 (1.5) | 12,611 (1.6) | 500 (0.5) |
| Edema (%) | FALSE | 887,613 (99.5) | 802,624 (99.5) | 95,636 (99.7) |
| | TRUE | 4,141 (0.5) | 3,886 (0.5) | 294 (0.3) |
| Chillbains (%) | FALSE | 891649 (100.0) | 806445 (100.0) | 95885 (100.0) |
| | TRUE | 105 (0.0) | 65 (0.0) | 45 (0.0) |
| Sepsis (%) | FALSE | 866,469 (97.2) | 781,276 (96.9) | 95,685 (99.7) |
| | TRUE | 25,285 (2.8) | 25,234 (3.1) | 245 (0.3) |
| Congestion (%) | FALSE | 884,446 (99.2) | 799,370 (99.1) | 95,745 (99.8) |
| | TRUE | 7,308 (0.8) | 7,140 (0.9) | 185 (0.2) |
| Dyspnea (%) | FALSE | 803,759 (90.1) | 721,135 (89.4) | 92,578 (96.5) |
| | TRUE | 87,995 (9.9) | 85,375 (10.6) | 3,352 (3.5) |
| Upper Respiratory Infection (%) | FALSE | 826,547 (92.7) | 742,953 (92.1) | 94,189 (98.2) |
| | TRUE | 65,207 (7.3) | 63,557 (7.9) | 1,741 (1.8) |
| Acute Respiratory Failure (%) | FALSE | 856,919 (96.1) | 771,789 (95.7) | 95,494 (99.5) |
| | TRUE | 34,835 (3.9) | 34,721 (4.3) | 436 (0.5) |
| Cough (%) | FALSE | 773,498 (86.7) | 691,288 (85.7) | 92,362 (96.3) |
| | TRUE | 118,256 (13.3) | 115,222 (14.3) | 3,568 (3.7) |
| Bronchitis (%) | FALSE | 879,904 (98.7) | 795,044 (98.6) | 95,538 (99.6) |
| | TRUE | 11,850 (1.3) | 11,466 (1.4) | 392 (0.4) |
| Bronchiolitis (%) | FALSE | 891,257 (99.9) | 806,018 (99.9) | 95920 (100.0) |
| | TRUE | 497 (0.1) | 492 (0.1) | 10 (0.0) |
| Pneumonia Full (%) | FALSE | 845,305 (94.8) | 760,583 (94.3) | 94,847 (98.9) |
| | TRUE | 46,449 (5.2) | 45,927 (5.7) | 1,083 (1.1) |
| Pneumonia Nos Or Sars (%) | FALSE | 846,991 (95.0) | 762,242 (94.5) | 94,893 (98.9) |
| | TRUE | 44,763 (5.0) | 44,268 (5.5) | 1,037 (1.1) |
| Lower Respiratory Nos (%) | FALSE | 887,880 (99.6) | 802,702 (99.5) | 95,855 (99.9) |
| | TRUE | 3,874 (0.4) | 3,808 (0.5) | 75 (0.1) |
| Acute Respiratory Distress Syndrome (%) | FALSE | 888,985 (99.7) | 803,755 (99.7) | 95,859 (99.9) |

| | | | | |
|---|---|---|---|---|
| | TRUE | 2,769 (0.3) | 2,755 (0.3) | 71 (0.1) |
| Other Specified Respiratory Disorders (%) | FALSE | 880,882 (98.8) | 796,390 (98.7) | 94,737 (98.8) |
| | TRUE | 10,872 (1.2) | 10,120 (1.3) | 1,193 (1.2) |
| Syncope (%) | FALSE | 883,380 (99.1) | 798,357 (99.0) | 95,675 (99.7) |
| | TRUE | 8,374 (0.9) | 8,153 (1.0) | 255 (0.3) |
| Chest Pain (%) | FALSE | 862,008 (96.7) | 777,452 (96.4) | 95,027 (99.1) |
| | TRUE | 29,746 (3.3) | 29,058 (3.6) | 903 (0.9) |
| Hyperlipidemia (%) | FALSE | 827,110 (92.8) | 747,718 (92.7) | 89,578 (93.4) |
| | TRUE | 64,644 (7.2) | 58,792 (7.3) | 6,352 (6.6) |
| Hemorrhage (%) | FALSE | 888,742 (99.7) | 803,582 (99.6) | 95,819 (99.9) |
| | TRUE | 3,012 (0.3) | 2,928 (0.4) | 111 (0.1) |
| Disseminated Intravascular Coagulation (%) | FALSE | 891481 (100.0) | 806239 (100.0) | 95926 (100.0) |
| | TRUE | 273 (0.0) | 271 (0.0) | 4 (0.0) |
| Thrombocytopenia (%) | FALSE | 882,836 (99.0) | 797,819 (98.9) | 95,605 (99.7) |
| | TRUE | 8,918 (1.0) | 8,691 (1.1) | 325 (0.3) |
| Embolism and Thrombosis (%) | FALSE | 882,355 (98.9) | 797,494 (98.9) | 95,363 (99.4) |
| | TRUE | 9,399 (1.1) | 9,016 (1.1) | 567 (0.6) |
| Stroke (%) | FALSE | 874,524 (98.1) | 790,001 (98.0) | 95,067 (99.1) |
| | TRUE | 17,230 (1.9) | 16,509 (2.0) | 863 (0.9) |
| Heart Failure (%) | FALSE | 854,090 (95.8) | 769,472 (95.4) | 95,088 (99.1) |
| | TRUE | 37,664 (4.2) | 37,038 (4.6) | 842 (0.9) |
| Diarrhea (%) | FALSE | 868,961 (97.4) | 784,161 (97.2) | 95,409 (99.5) |
| | TRUE | 22,793 (2.6) | 22,349 (2.8) | 521 (0.5) |
| Nausea (%) | FALSE | 865,016 (97.0) | 780,158 (96.7) | 95,493 (99.5) |
| | TRUE | 26,738 (3.0) | 26,352 (3.3) | 437 (0.5) |
| Vomiting (%) | FALSE | 874,434 (98.1) | 789,364 (97.9) | 95,724 (99.8) |
| | TRUE | 17,320 (1.9) | 17,146 (2.1) | 206 (0.2) |
| Abdominal Pain (%) | FALSE | 856,932 (96.1) | 773,178 (95.9) | 94,191 (98.2) |
| | TRUE | 34,822 (3.9) | 33,332 (4.1) | 1,739 (1.8) |
| Dehydration (%) | FALSE | 876,357 (98.3) | 791,185 (98.1) | 95,776 (99.8) |
| | TRUE | 15,397 (1.7) | 15,325 (1.9) | 154 (0.2) |
| Loss of Appetite (%) | FALSE | 889,407 (99.7) | 804,220 (99.7) | 95,847 (99.9) |
| | TRUE | 2,347 (0.3) | 2,290 (0.3) | 83 (0.1) |
| Gerd (%) | FALSE | 845,551 (94.8) | 763,464 (94.7) | 92,488 (96.4) |
| | TRUE | 46,203 (5.2) | 43,046 (5.3) | 3,442 (3.6) |
| Acute Kidney Failure (%) | FALSE | 857,104 (96.1) | 772,005 (95.7) | 95,577 (99.6) |
| | TRUE | 34,650 (3.9) | 34,505 (4.3) | 353 (0.4) |
| Headache (%) | FALSE | 869,704 (97.5) | 785,010 (97.3) | 95,288 (99.3) |
| | TRUE | 22,050 (2.5) | 21,500 (2.7) | 642 (0.7) |

| | | | | |
|---|---|---|---|---|
| Confusion or Disorientation (%) | FALSE | 877,492 (98.4) | 792,309 (98.2) | 95,790 (99.9) |
| | TRUE | 14,262 (1.6) | 14,201 (1.8) | 140 (0.1) |
| Disturbances of Smell Or Taste (%) | FALSE | 888,385 (99.6) | 803,394 (99.6) | 95,635 (99.7) |
| | TRUE | 3,369 (0.4) | 3,116 (0.4) | 295 (0.3) |
| Hypersomnia (%) | FALSE | 891313 (100.0) | 806131 (100.0) | 95,860 (99.9) |
| | TRUE | 441 (0.0) | 379 (0.0) | 70 (0.1) |
| Dizziness (%) | FALSE | 880,295 (98.7) | 795,788 (98.7) | 95,123 (99.2) |
| | TRUE | 11,459 (1.3) | 10,722 (1.3) | 807 (0.8) |
| Conjunctivitis (%) | FALSE | 890,440 (99.9) | 805,353 (99.9) | 95,730 (99.8) |
| | TRUE | 1,314 (0.1) | 1,157 (0.1) | 200 (0.2) |
| Visual Discomfort (%) | FALSE | 891610 (100.0) | 806370 (100.0) | 95924 (100.0) |
| | TRUE | 144 (0.0) | 140 (0.0) | 6 (0.0) |
| Edema of Eye (%) | FALSE | 891705 (100.0) | 806465 (100.0) | 95924 (100.0) |
| | TRUE | 49 (0.0) | 45 (0.0) | 6 (0.0) |
| Other General Signs or Symptoms (%) | FALSE | 813,100 (91.2) | 729,969 (90.5) | 92,839 (96.8) |
| | TRUE | 78,654 (8.8) | 76,541 (9.5) | 3,091 (3.2) |
| COVID_related U codes (%) | FALSE | 845,508 (94.8) | 763,872 (94.7) | 89,614 (93.4) |
| | TRUE | 46,246 (5.2) | 42,638 (5.3) | 6,316 (6.6) |
| COVID-related B codes (%) | FALSE | 885,522 (99.3) | 800,304 (99.2) | 95905 (100.0) |
| | TRUE | 6,232 (0.7) | 6,206 (0.8) | 25 (0.0) |
| Pre-Procedural Codes (%) | FALSE | 789,122 (88.5) | 704,939 (87.4) | 94,671 (98.7) |
| | TRUE | 102,632 (11.5) | 101,571 (12.6) | 1,259 (1.3) |
| Screening and Exposure (%) | FALSE | 498,226 (55.9) | 446,620 (55.4) | 55,305 (57.7) |
| | TRUE | 393,528 (44.1) | 359,890 (44.6) | 40,625 (42.3) |
| Screening and Exposure Antibody (%) | FALSE | 878,688 (98.5) | 806146 (100.0) | 82,319 (85.8) |
| | TRUE | 13,066 (1.5) | 364 (0.0) | 13,611 (14.2) |

Supplement H. Distribution of Comorbidities/ Existing Conditions by Cohort

| **Variable** [a] | **Level** | **Cohort 1** | **Cohort 2** | **Cohort 3** |
|---|---|---|---|---|
| n | | 891,754 | 806,510 | 95,930 |
| Charlson Comorbidity Score (%) | 0 | 386,110 (43.3) | 340,416 (42.2) | 51,374 (53.6) |
| | 1 | 165,058 (18.5) | 146,543 (18.2) | 20,547 (21.4) |
| | 2 | 82,845 (9.3) | 75,803 (9.4) | 8,080 (8.4) |
| | 3 | 49,290 (5.5) | 46,174 (5.7) | 3,691 (3.8) |
| | 4 | 31,553 (3.5) | 30,211 (3.7) | 1,677 (1.7) |
| | 5 | 20,945 (2.3) | 20,294 (2.5) | 844 (0.9) |
| | 6 | 13,350 (1.5) | 13,038 (1.6) | 440 (0.5) |
| | 7+ | 14,485 (1.6) | 14,290 (1.8) | 321 (0.3) |
| | Insufficient (≤365) days of database history | 128,118 (14.4) | 119,741 (14.8) | 8,956 (9.3) |
| Smoking Status (%) | Never smoked | 377,589 (42.3) | 333,780 (41.4) | 50,856 (53.0) |
| | Not currently smoking | 24,255 (2.7) | 21,363 (2.6) | 2,265 (2.4) |
| | Previously smoked | 193,044 (21.6) | 173,714 (21.5) | 21,506 (22.4) |
| | Current smoker | 117,800 (13.2) | 112,327 (13.9) | 5,564 (5.8) |
| | Unknown | 179,066 (20.1) | 165,326 (20.5) | 15,739 (16.4) |
| BMI (%) | Normal | 129,668 (14.5) | 114,657 (14.2) | 17,576 (18.3) |
| | Underweight | 25,684 (2.9) | 23,974 (3.0) | 1,981 (2.1) |
| | Overweight | 235,020 (26.4) | 207,080 (25.7) | 31,326 (32.7) |
| | Obese | 280,185 (31.4) | 254,521 (31.6) | 28,055 (29.2) |
| | Unknown | 221,197 (24.8) | 206,278 (25.6) | 16,992 (17.7) |
| CCI: Myocardial infarction (%) | FALSE | 721,868 (80.9) | 646,291 (80.1) | 85,275 (88.9) |
| | TRUE | 41,768 (4.7) | 40,478 (5.0) | 1,699 (1.8) |
| | Insufficient (≤365) days of database history | 128,118 (14.4) | 119,741 (14.8) | 8,956 (9.3) |
| CCI: Congestive heart failure (%) | FALSE | 692,182 (77.6) | 617,734 (76.6) | 84,028 (87.6) |
| | TRUE | 71,454 (8.0) | 69,035 (8.6) | 2,946 (3.1) |
| | Insufficient (≤365) days of database history | 128,118 (14.4) | 119,741 (14.8) | 8,956 (9.3) |

| | | | | |
|---|---|---|---|---|
| CCI: Peripheral vascular disease (%) | FALSE | 705,484 (79.1) | 631,688 (78.3) | 83,290 (86.8) |
| | TRUE | 58,152 (6.5) | 55,081 (6.8) | 3,684 (3.8) |
| | Insufficient (≤365) days of database history | 128,118 (14.4) | 119,741 (14.8) | 8,956 (9.3) |
| CCI: Cerebrovascular disease (%) | FALSE | 710,626 (79.7) | 636,989 (79.0) | 83,302 (86.8) |
| | TRUE | 53,010 (5.9) | 49,780 (6.2) | 3,672 (3.8) |
| | Insufficient (≤365) days of database history | 128,118 (14.4) | 119,741 (14.8) | 8,956 (9.3) |
| CCI: Dementia (%) | FALSE | 740,264 (83.0) | 663,868 (82.3) | 86,448 (90.1) |
| | TRUE | 23,372 (2.6) | 22,901 (2.8) | 526 (0.5) |
| | Insufficient (≤365) days of database history | 128,118 (14.4) | 119,741 (14.8) | 8,956 (9.3) |
| CCI: Chronic pulmonary disease (%) | FALSE | 594,589 (66.7) | 529,659 (65.7) | 73,138 (76.2) |
| | TRUE | 169,047 (19.0) | 157,110 (19.5) | 13,836 (14.4) |
| | Insufficient (≤365) days of database history | 128,118 (14.4) | 119,741 (14.8) | 8,956 (9.3) |
| CCI: Rheumatic disease (%) | FALSE | 741,491 (83.1) | 666,795 (82.7) | 84,523 (88.1) |
| | TRUE | 22,145 (2.5) | 19,974 (2.5) | 2,451 (2.6) |
| | Insufficient (≤365) days of database history | 128,118 (14.4) | 119,741 (14.8) | 8,956 (9.3) |
| CCI: Peptic ulcer disease (%) | FALSE | 753,195 (84.5) | 676,825 (83.9) | 86,349 (90.0) |
| | TRUE | 10,441 (1.2) | 9,944 (1.2) | 625 (0.7) |
| | Insufficient (≤365) days of database history | 128,118 (14.4) | 119,741 (14.8) | 8,956 (9.3) |
| CCI: Mild liver disease (%) | FALSE | 719,323 (80.7) | 645,438 (80.0) | 83,406 (86.9) |
| | TRUE | 44,313 (5.0) | 41,331 (5.1) | 3,568 (3.7) |
| | Insufficient (≤365) days of database history | 128,118 (14.4) | 119,741 (14.8) | 8,956 (9.3) |
| CCI: Diabetes without chronic complications (%) | FALSE | 640,231 (71.8) | 573,025 (71.0) | 76,325 (79.6) |
| | TRUE | 123,405 (13.8) | 113,744 (14.1) | 10,649 (11.1) |
| | Insufficient (≤365) days of database history | 128,118 (14.4) | 119,741 (14.8) | 8,956 (9.3) |
| CCI: Diabetes with chronic complications (%) | FALSE | 699,026 (78.4) | 625,278 (77.5) | 83,430 (87.0) |
| | TRUE | 64,610 (7.2) | 61,491 (7.6) | 3,544 (3.7) |
| | Insufficient (≤365) days of database history | 128,118 (14.4) | 119,741 (14.8) | 8,956 (9.3) |

| | | | | |
|---|---|---|---|---|
| CCI: Hemiplegia or paraplegia (%) | FALSE | 754,004 (84.6) | 677,356 (84.0) | 86,688 (90.4) |
| | TRUE | 9,632 (1.1) | 9,413 (1.2) | 286 (0.3) |
| | Insufficient (≤365) days of database history | 128,118 (14.4) | 119,741 (14.8) | 8,956 (9.3) |
| CCI: Renal disease (%) | FALSE | 688,086 (77.2) | 614,596 (76.2) | 82,910 (86.4) |
| | TRUE | 75,550 (8.5) | 72,173 (8.9) | 4,064 (4.2) |
| | Insufficient (≤365) days of database history | 128,118 (14.4) | 119,741 (14.8) | 8,956 (9.3) |
| CCI: Any malignancy (except skin) (%) | FALSE | 656,345 (73.6) | 587,171 (72.8) | 77,385 (80.7) |
| | TRUE | 107,291 (12.0) | 99,598 (12.3) | 9,589 (10.0) |
| | Insufficient (≤365) days of database history | 128,118 (14.4) | 119,741 (14.8) | 8,956 (9.3) |
| CCI: Moderate or severe liver disease (%) | FALSE | 757,804 (85.0) | 681,080 (84.4) | 86,777 (90.5) |
| | TRUE | 5,832 (0.7) | 5,689 (0.7) | 197 (0.2) |
| | Insufficient (≤365) days of database history | 128,118 (14.4) | 119,741 (14.8) | 8,956 (9.3) |
| CCI: Metastatic solid tumor (%) | FALSE | 745,518 (83.6) | 669,569 (83.0) | 85,317 (88.9) |
| | TRUE | 18,118 (2.0) | 17,200 (2.1) | 1,657 (1.7) |
| | Insufficient (≤365) days of database history | 128,118 (14.4) | 119,741 (14.8) | 8,956 (9.3) |
| CCI: AIDS/HIV (%) | FALSE | 760,555 (85.3) | 684,388 (84.9) | 86,195 (89.9) |
| | TRUE | 3,081 (0.3) | 2,381 (0.3) | 779 (0.8) |
| | Insufficient (≤365) days of database history | 128,118 (14.4) | 119,741 (14.8) | 8,956 (9.3) |
| Non-CCI: Asthma (%) | FALSE | 675,098 (75.7) | 605,259 (75.0) | 78,685 (82.0) |
| | TRUE | 88,538 (9.9) | 81,510 (10.1) | 8,289 (8.6) |
| | Insufficient (≤365) days of database history | 128,118 (14.4) | 119,741 (14.8) | 8,956 (9.3) |
| Non-CCI: Hypertension (%) | FALSE | 470,867 (52.8) | 421,729 (52.3) | 56,548 (58.9) |
| | TRUE | 292,769 (32.8) | 265,040 (32.9) | 30,426 (31.7) |
| | Insufficient (≤365) days of database history | 128,118 (14.4) | 119,741 (14.8) | 8,956 (9.3) |
| Non-CCI: Hyperlipidemia (%) | FALSE | 505,470 (56.7) | 460,529 (57.1) | 52,072 (54.3) |
| | TRUE | 258,166 (29.0) | 226,240 (28.1) | 34,902 (36.4) |
| | Insufficient (≤365) days of database history | 128,118 (14.4) | 119,741 (14.8) | 8,956 (9.3) |
| Non-CCI: Pregnancy (%) | FALSE | 763,733 (85.6) | 685,973 (85.1) | 88,025 (91.8) |

| | | | | |
|---|---|---|---|---|
| | TRUE | 24,112 (2.7) | 23,175 (2.9) | 1,103 (1.1) |
| | Insufficient (≤365) days of database history | 103,909 (11.7) | 97,362 (12.1) | 6,802 (7.1) |

BMI=body mass index; CCI=Charlson Comorbidity Index; Non-CCI=Not derived as part of the CCI.

[a] Patients with ≤365 days of database history were not considered for comorbidities based on ICD codes

Supplement I. Cohort 2 Pediatric Population: SARS-CoV-2 Positivity Rates by Sociodemographic and Clinical Characteristics

| Variable | Level | Positivity (%) | Test Result | | | | |
|---|---|---|---|---|---|---|---|
| | | | Overall | Positive | Negative | Uninterpretable | None[a] |
| n | | 5.7 | 39,691 | 2,120 | 35,121 | 580 | 1,870 |
| Age [min, Q1, median, Q3, max] [b] | | NA | [0, 2, 7, 14, 17] | [0, 3, 10, 15, 17] | [0, 2, 7, 13, 17] | [0, 2, 6, 12.25, 17] | [0, 2, 5.5, 12, 17] |
| Age (%) | 0-4 | 4.7 | 15,069 (38.0) | 657 (31.0) | 13,357 (38.0) | 235 (40.5) | 820 (43.9) |
| | 5-17 | 6.3 | 24,622 (62.0) | 1,463 (69.0) | 21,764 (62.0) | 345 (59.5) | 1,050 (56.1) |
| Sex (%) | Female | 5.6 | 19,271 (48.6) | 1,020 (48.1) | 17,073 (48.6) | 296 (51.0) | 882 (47.2) |
| | Male | 5.7 | 20,360 (51.3) | 1,098 (51.8) | 17,998 (51.2) | 282 (48.6) | 982 (52.5) |
| | Unknown | 3.8 | 60 (0.2) | 2 (0.1) | 50 (0.1) | 2 (0.3) | 6 (0.3) |
| Race (%) | African American | 9.6 | 4,067 (10.2) | 357 (16.8) | 3,348 (9.5) | 58 (10.0) | 304 (16.3) |
| | Asian | 7.4 | 848 (2.1) | 61 (2.9) | 761 (2.2) | 5 (0.9) | 21 (1.1) |
| | Caucasian | 3.4 | 24,170 (60.9) | 775 (36.6) | 21,843 (62.2) | 411 (70.9) | 1,141 (61.0) |
| | Unknown | 9.2 | 10,606 (26.7) | 927 (43.7) | 9,169 (26.1) | 106 (18.3) | 404 (21.6) |
| Ethnicity (%) | Hispanic | 14.4 | 5,164 (13.0) | 712 (33.6) | 4,238 (12.1) | 71 (12.2) | 143 (7.6) |
| | Not Hispanic | 3.9 | 27,118 (68.3) | 992 (46.8) | 24,472 (69.7) | 432 (74.5) | 1,222 (65.3) |
| | Unknown | 6.1 | 7,409 (18.7) | 416 (19.6) | 6,411 (18.3) | 77 (13.3) | 505 (27.0) |
| Race/Ethnicity (%) | Non-Hispanic white | 2.6 | 20,512 (51.7) | 500 (23.6) | 18,774 (53.5) | 357 (61.6) | 881 (47.1) |
| | Asian | 7.5 | 757 (1.9) | 55 (2.6) | 679 (1.9) | 5 (0.9) | 18 (1.0) |
| | Hispanic | 12.7 | 1,779 (4.5) | 211 (10.0) | 1,455 (4.1) | 35 (6.0) | 78 (4.2) |
| | Non-Hispanic black | 8.9 | 3,527 (8.9) | 288 (13.6) | 2,935 (8.4) | 47 (8.1) | 257 (13.7) |
| | Unknown | 8.6 | 13,116 (33.0) | 1,066 (50.3) | 11,278 (32.1) | 136 (23.4) | 636 (34.0) |
| Region (%) | Midwest | 5.0 | 19,754 (49.8) | 903 (42.6) | 17,040 (48.5) | 373 (64.3) | 1,438 (76.9) |
| | Northeast | 8.4 | 9,579 (24.1) | 780 (36.8) | 8,539 (24.3) | 100 (17.2) | 160 (8.6) |
| | South | 5.2 | 5,739 (14.5) | 290 (13.7) | 5,303 (15.1) | 31 (5.3) | 115 (6.1) |
| | West | 2.9 | 3,645 (9.2) | 101 (4.8) | 3,374 (9.6) | 52 (9.0) | 118 (6.3) |
| | Unknown | 5.0 | 974 (2.5) | 46 (2.2) | 865 (2.5) | 24 (4.1) | 39 (2.1) |

| | | | | | | | |
|---|---|---|---|---|---|---|---|
| Insurance Type (%) | Commercial | 4.9 | 22,114 (55.7) | 1,022 (48.2) | 19,765 (56.3) | 351 (60.5) | 976 (52.2) |
| | Medicaid | 7.5 | 10,966 (27.6) | 771 (36.4) | 9,461 (26.9) | 106 (18.3) | 628 (33.6) |
| | Medicare | 4.2 | 52 (0.1) | 2 (0.1) | 46 (0.1) | 2 (0.3) | 2 (0.1) |
| | Other Payor type | 4.8 | 3,301 (8.3) | 147 (6.9) | 2,945 (8.4) | 76 (13.1) | 133 (7.1) |
| | Uninsured | 3.2 | 154 (0.4) | 4 (0.2) | 121 (0.3) | 8 (1.4) | 21 (1.1) |
| | Unknown | 5.9 | 3,104 (7.8) | 174 (8.2) | 2,783 (7.9) | 37 (6.4) | 110 (5.9) |
| Index Setting (%) | Ambulatory | 5.9 | 15,298 (38.5) | 852 (40.2) | 13,473 (38.4) | 119 (20.5) | 854 (45.7) |
| | Inpatient | 5.7 | 12,518 (31.5) | 668 (31.5) | 11,037 (31.4) | 177 (30.5) | 636 (34.0) |
| | Lab | 4 | 4,423 (11.1) | 164 (7.7) | 3,963 (11.3) | 238 (41.0) | 58 (3.1) |
| | Other | 7.7 | 4,703 (11.8) | 353 (16.7) | 4,221 (12.0) | 25 (4.3) | 104 (5.6) |
| | Unknown | 3.3 | 2,749 (6.9) | 83 (3.9) | 2,427 (6.9) | 21 (3.6) | 218 (11.7) |
| Test source (viral) (%) | Upper respiratory | 3.6 | 5,117 (12.9) | 178 (8.4) | 4,826 (13.7) | 113 (19.5) | 0 (0.0) |
| | Lower respiratory | 0.0 | 1 (0.0) | 0 (0.0) | 1 (0.0) | 0 (0.0) | 0 (0.0) |
| | Unknown | 6.0 | 34,573 (87.1) | 1,942 (91.6) | 30,294 (86.3) | 467 (80.5) | 1870 (100.0) |
| Positive for any seasonal respiratory isolate (%) | At least 1 test positive | 1.7 | 59 (0.1) | 1 (0.0) | 57 (0.2) | 1 (0.2) | 0 (0.0) |
| | Not tested | 5.8 | 34,237 (86.3) | 1,869 (88.2) | 30,284 (86.2) | 416 (71.7) | 1,668 (89.2) |
| | Tested but no positives | 5.0 | 5,395 (13.6) | 250 (11.8) | 4,780 (13.6) | 163 (28.1) | 202 (10.8) |
| Days since COVID-like illness (%) | No COVID-like illness observed | 4.5 | 31,877 (80.3) | 1,344 (63.4) | 28,467 (81.1) | 468 (80.7) | 1,598 (85.5) |
| | 0 | 12.0 | 5,765 (14.5) | 660 (31.1) | 4,833 (13.8) | 83 (14.3) | 189 (10.1) |
| | 1 - 3 | 8.8 | 1,004 (2.5) | 85 (4.0) | 881 (2.5) | 9 (1.6) | 29 (1.6) |
| | 4 - 7 | 5.7 | 203 (0.5) | 11 (0.5) | 181 (0.5) | 2 (0.3) | 9 (0.5) |
| | 8 - 14 | 3.0 | 178 (0.4) | 5 (0.2) | 160 (0.5) | 6 (1.0) | 7 (0.4) |
| | 15 - 21 | 3.2 | 166 (0.4) | 5 (0.2) | 151 (0.4) | 1 (0.2) | 9 (0.5) |
| | 22 - 28 | 2.3 | 144 (0.4) | 3 (0.1) | 127 (0.4) | 4 (0.7) | 10 (0.5) |
| | 29 - 35 | 3.5 | 154 (0.4) | 5 (0.2) | 136 (0.4) | 4 (0.7) | 9 (0.5) |
| | 36 - 42 | 1.1 | 200 (0.5) | 2 (0.1) | 185 (0.5) | 3 (0.5) | 10 (0.5) |
| Number of symptoms (%)[c] | 0 | 4.8 | 22,328 (56.3) | 1,001 (47.2) | 19,846 (56.5) | 330 (56.9) | 1,151 (61.6) |
| | 1-3 | 6.9 | 16,640 (41.9) | 1,076 (50.8) | 14,627 (41.6) | 229 (39.5) | 708 (37.9) |

| | | | | | | | |
|---|---|---|---|---|---|---|---|
| | 4-7 | 6.2 | 721 (1.8) | 43 (2.0) | 646 (1.8) | 21 (3.6) | 11 (0.6) |
| | 8-11 | 0.0 | 2 (0.0) | 0 (0.0) | 2 (0.0) | 0 (0.0) | 0 (0.0) |
| | 12+ | NaN | 0 (0.0) | 0 (0.0) | 0 (0.0) | 0 (0.0) | 0 (0.0) |
| Charlson Comorbidity Score (%) | 0 | 5.8 | 21,897 (55.2) | 1,202 (56.7) | 19,463 (55.4) | 332 (57.2) | 900 (48.1) |
| | 1 | 5.0 | 4,863 (12.3) | 227 (10.7) | 4,296 (12.2) | 82 (14.1) | 258 (13.8) |
| | 2 | 2.3 | 892 (2.2) | 19 (0.9) | 807 (2.3) | 20 (3.4) | 46 (2.5) |
| | 3 | 2.9 | 229 (0.6) | 6 (0.3) | 199 (0.6) | 1 (0.2) | 23 (1.2) |
| | 4 | 1.8 | 62 (0.2) | 1 (0.0) | 56 (0.2) | 1 (0.2) | 4 (0.2) |
| | 5 | 0.0 | 18 (0.0) | 0 (0.0) | 16 (0.0) | 0 (0.0) | 2 (0.1) |
| | 6 | 0.0 | 6 (0.0) | 0 (0.0) | 6 (0.0) | 0 (0.0) | 0 (0.0) |
| | 7+ | NaN | 0 (0.0) | 0 (0.0) | 0 (0.0) | 0 (0.0) | 0 (0.0) |
| | Insufficient (≤365) days of database history [d] | 6.1 | 11,724 (29.5) | 665 (31.4) | 10,278 (29.3) | 144 (24.8) | 637 (34.1) |
| BMI (%) | Normal | 5.2 | 8,225 (20.7) | 407 (19.2) | 7,363 (21.0) | 122 (21.0) | 333 (17.8) |
| | Underweight | 4.3 | 13,524 (34.1) | 542 (25.6) | 12,118 (34.5) | 199 (34.3) | 665 (35.6) |
| | Overweight | 6.9 | 3,157 (8.0) | 206 (9.7) | 2,795 (8.0) | 56 (9.7) | 100 (5.3) |
| | Obese | 8.5 | 2,313 (5.8) | 188 (8.9) | 2,018 (5.7) | 31 (5.3) | 76 (4.1) |
| | Unknown | 6.7 | 12,472 (31.4) | 777 (36.7) | 10,827 (30.8) | 172 (29.7) | 696 (37.2) |

BMI=body mass index; IQR=interquartile range

[a] indexed on procedural code no test result available

[b] 0, 25th, 50th, 75th, and 100th percentiles.

[c] Symptoms were chosen as those associated with positivity in C2 modeling.

[d] Patients with ≤365 days of database history were not considered for comorbidities.

Supplement J. Cohort 3 Pediatric Population: SARS-CoV-2 Positivity Rates by Sociodemographic and Clinical Characteristics

| Variable | Level | Positivity (%) | Test Result | | | | |
|---|---|---|---|---|---|---|---|
| | | | Overall | Positive | Negative | Uninterp retable | None[a] |
| n | | 17.4 | 2,962 | 490 | 2,331 | 35 | 106 |
| Age [min, Q1, median, Q3, max] [b] | | NA | [0, 6, 11, 15, 17] | [0, 5.25, 11, 15, 17] | [0, 6, 11, 15, 17] | [2, 8, 11, 15, 17] | [0, 3, 9, 13.75, 17] |
| Age (%) | 0-4 | 18.9 | 576 (19.4) | 102 (20.8) | 439 (18.8) | 3 (8.6) | 32 (30.2) |
| | 5-17 | 17.0 | 2,386 (80.6) | 388 (79.2) | 1,892 (81.2) | 32 (91.4) | 74 (69.8) |
| Sex (%) | Female | 16.7 | 1,468 (49.6) | 233 (47.6) | 1,163 (49.9) | 18 (51.4) | 54 (50.9) |
| | Male | 18.1 | 1,492 (50.4) | 257 (52.4) | 1,166 (50.0) | 17 (48.6) | 52 (49.1) |
| | Unknown | 0.0 | 2 (0.1) | 0 (0.0) | 2 (0.1) | 0 (0.0) | 0 (0.0) |
| Race (%) | African American | 24.0 | 159 (5.4) | 36 (7.3) | 114 (4.9) | 1 (2.9) | 8 (7.5) |
| | Asian | 22.5 | 76 (2.6) | 16 (3.3) | 55 (2.4) | 2 (5.7) | 3 (2.8) |
| | Caucasian | 14.8 | 1,769 (59.7) | 249 (50.8) | 1,429 (61.3) | 16 (45.7) | 75 (70.8) |
| | Unknown | 20.5 | 958 (32.3) | 189 (38.6) | 733 (31.4) | 16 (45.7) | 20 (18.9) |
| Ethnicity (%) | Hispanic | 23.0 | 152 (5.1) | 31 (6.3) | 104 (4.5) | 3 (8.6) | 14 (13.2) |
| | Not Hispanic | 10.2 | 1,589 (53.6) | 153 (31.2) | 1,344 (57.7) | 13 (37.1) | 79 (74.5) |
| | Unknown | 25.7 | 1,221 (41.2) | 306 (62.4) | 883 (37.9) | 19 (54.3) | 13 (12.3) |
| Race/Ethnicity (%) | Non-Hispanic white | 8.5 | 1,301 (43.9) | 104 (21.2) | 1,119 (48.0) | 11 (31.4) | 67 (63.2) |
| | Asian | 17.9 | 61 (2.1) | 10 (2.0) | 46 (2.0) | 2 (5.7) | 3 (2.8) |
| | Hispanic | 24.3 | 76 (2.6) | 17 (3.5) | 53 (2.3) | 1 (2.9) | 5 (4.7) |
| | Non-Hispanic black | 23.1 | 113 (3.8) | 25 (5.1) | 83 (3.6) | 0 (0.0) | 5 (4.7) |
| | Unknown | 24.5 | 1,411 (47.6) | 334 (68.2) | 1,030 (44.2) | 21 (60.0) | 26 (24.5) |
| Region (%) | Midwest | 4.8 | 831 (28.1) | 38 (7.8) | 760 (32.6) | 1 (2.9) | 32 (30.2) |
| | Northeast | 25.2 | 1,614 (54.5) | 382 (78.0) | 1,134 (48.6) | 25 (71.4) | 73 (68.9) |
| | South | 12.6 | 354 (12.0) | 44 (9.0) | 306 (13.1) | 4 (11.4) | 0 (0.0) |
| | West | 7.4 | 29 (1.0) | 2 (0.4) | 25 (1.1) | 2 (5.7) | 0 (0.0) |
| | Unknown | 18.5 | 134 (4.5) | 24 (4.9) | 106 (4.5) | 3 (8.6) | 1 (0.9) |
| Insurance Type (%) | Commercial | 12.2 | 1,871 (63.2) | 216 (44.1) | 1,550 (66.5) | 19 (54.3) | 86 (81.1) |
| | Medicaid | 40.2 | 454 (15.3) | 173 (35.3) | 257 (11.0) | 7 (20.0) | 17 (16.0) |

| | | | | | | | |
|---|---|---|---|---|---|---|---|
| | Other Payor type | 4.9 | 126 (4.3) | 6 (1.2) | 116 (5.0) | 1 (2.9) | 3 (2.8) |
| | Unknown | 18.9 | 511 (17.3) | 95 (19.4) | 408 (17.5) | 8 (22.9) | 0 (0.0) |
| Index Setting (%) | Ambulatory | 17.6 | 1,447 (48.9) | 246 (50.2) | 1,153 (49.5) | 5 (14.3) | 43 (40.6) |
| | Inpatient | 16.6 | 371 (12.5) | 55 (11.2) | 277 (11.9) | 0 (0.0) | 39 (36.8) |
| | Lab | 15.8 | 474 (16.0) | 72 (14.7) | 385 (16.5) | 0 (0.0) | 17 (16.0) |
| | Other | 20.4 | 281 (9.5) | 55 (11.2) | 214 (9.2) | 7 (20.0) | 5 (4.7) |
| | Unknown | 17.0 | 389 (13.1) | 62 (12.7) | 302 (13.0) | 23 (65.7) | 2 (1.9) |
| Positive for any seasonal respiratory isolate (%) | At least 1 test positive | 25.0 | 4 (0.1) | 1 (0.2) | 3 (0.1) | 0 (0.0) | 0 (0.0) |
| | Not tested | 17.4 | 2,684 (90.6) | 447 (91.2) | 2,118 (90.9) | 35 (100.0) | 84 (79.2) |
| | Tested but no positives | 16.7 | 274 (9.3) | 42 (8.6) | 210 (9.0) | 0 (0.0) | 22 (20.8) |
| Days since COVID-like illness (%) | No COVID-like illness observed | 16.8 | 2,637 (89.0) | 426 (86.9) | 2,103 (90.2) | 30 (85.7) | 78 (73.6) |
| | 0 | 18.9 | 171 (5.8) | 30 (6.1) | 129 (5.5) | 3 (8.6) | 9 (8.5) |
| | 1 - 3 | 24.4 | 51 (1.7) | 11 (2.2) | 34 (1.5) | 1 (2.9) | 5 (4.7) |
| | 4 - 7 | 11.5 | 30 (1.0) | 3 (0.6) | 23 (1.0) | 0 (0.0) | 4 (3.8) |
| | 8 - 14 | 36.8 | 22 (0.7) | 7 (1.4) | 12 (0.5) | 0 (0.0) | 3 (2.8) |
| | 15 - 21 | 50.0 | 12 (0.4) | 4 (0.8) | 4 (0.2) | 1 (2.9) | 3 (2.8) |
| | 22 - 28 | 33.3 | 16 (0.5) | 5 (1.0) | 10 (0.4) | 0 (0.0) | 1 (0.9) |
| | 29 - 35 | 27.3 | 13 (0.4) | 3 (0.6) | 8 (0.3) | 0 (0.0) | 2 (1.9) |
| | 36 - 42 | 11.1 | 10 (0.3) | 1 (0.2) | 8 (0.3) | 0 (0.0) | 1 (0.9) |
| Charlson Comorbidity Score (%) | 0 | 18.9 | 1,774 (59.9) | 319 (65.1) | 1,371 (58.8) | 20 (57.1) | 64 (60.4) |
| | 1 | 12.0 | 361 (12.2) | 40 (8.2) | 293 (12.6) | 6 (17.1) | 22 (20.8) |
| | 2 | 16.3 | 50 (1.7) | 8 (1.6) | 41 (1.8) | 0 (0.0) | 1 (0.9) |
| | 3 | 16.7 | 9 (0.3) | 1 (0.2) | 5 (0.2) | 0 (0.0) | 3 (2.8) |
| | 4 | 0.0 | 2 (0.1) | 0 (0.0) | 2 (0.1) | 0 (0.0) | 0 (0.0) |
| | 5 | NaN | 0 (0.0) | 0 (0.0) | 0 (0.0) | 0 (0.0) | 0 (0.0) |
| | 6 | 0.0 | 1 (0.0) | 0 (0.0) | 1 (0.0) | 0 (0.0) | 0 (0.0) |
| | 7+ | NaN | 0 (0.0) | 0 (0.0) | 0 (0.0) | 0 (0.0) | 0 (0.0) |
| | Insufficient (≤365) days of database history [c] | 16.5 | 765 (25.8) | 122 (24.9) | 618 (26.5) | 9 (25.7) | 16 (15.1) |
| BMI (%) | Normal | 19.3 | 736 (24.8) | 136 (27.8) | 569 (24.4) | 8 (22.9) | 23 (21.7) |
| | Underweight | 14.9 | 1,045 (35.3) | 148 (30.2) | 842 (36.1) | 11 (31.4) | 44 (41.5) |

| | | | | | | | |
|---|---|---|---|---|---|---|---|
| | Overweight | 22.0 | 275 (9.3) | 58 (11.8) | 206 (8.8) | 4 (11.4) | 7 (6.6) |
| | Obese | 23.2 | 110 (3.7) | 23 (4.7) | 76 (3.3) | 2 (5.7) | 9 (8.5) |
| | Unknown | 16.4 | 796 (26.9) | 125 (25.5) | 638 (27.4) | 10 (28.6) | 23 (21.7) |
| Max previous viral test results (positive > negative > unknown) (%) | Not tested | 17.2 | 2,501 (84.4) | 413 (84.3) | 1,994 (85.5) | 27 (77.1) | 67 (63.2) |
| | Non-definitive result | 0 | 9 (0.3) | 0 (0.0) | 9 (0.4) | 0 (0.0) | 0 (0.0) |
| | Negative | 14.6 | 420 (14.2) | 55 (11.2) | 322 (13.8) | 7 (20.0) | 36 (34.0) |
| | Positive | 78.6 | 32 (1.1) | 22 (4.5) | 6 (0.3) | 1 (2.9) | 3 (2.8) |

BMI=body mass index; IQR=interquartile range.
[a] indexed on procedural code no test result available
[b] 0, 25th, 50th, 75th, and 100th percentiles.
[c] Patients with ≤365 days of database history were not considered for comorbidities.

Supplement K. Concordance between SARS-CoV-2 Viral and Antibody Test Among Patients <18 years [a]

| | | | | Viral test | |
|---|---|---|---|---|---|
| **Weeks Since Viral Test** | **N Patients** | **Overall Concordance (%)** | **Antibody Test** | *N Negative (% of viral)* | *N Positive (% of viral)* |
| Overall | 405 | 84.7 | *Negative* | 323 (85.0%) | 5 (20.0%) |
| | | | *Positive* | 57 (15.0%) | 20 (80.0%) |

[a] Among patients who had definitive results for both tests and the viral test was the initial test.
Note: insufficient patient numbers restricted our assessment of concordance by time interval between tests.

Supplement L. Cohort 2: Odds Ratios and 95% Confidence Intervals Obtained from Multivariable Logistic Models (Corresponds to Forest Plot in Figure 4)
OR and 95% CI Cohort 2

| **Type** | **Term** | **OR (95% CI)** |
|---|---|---|
| Intercept | (Intercept) | 0.05 (0.04, 0.05) |
| Sociodemographic | Age - <18 | 0.60 (0.57, 0.64) |
| Sociodemographic | Age - 35-44 | 0.93 (0.90, 0.96) |
| Sociodemographic | Age - 45-54 | 1.07 (1.03, 1.10) |
| Sociodemographic | Age - 55-64 | 1.08 (1.05, 1.12) |
| Sociodemographic | Age - 65+ | 1.11 (1.06, 1.15) |
| Sociodemographic | Insurance Type - Medicaid | 1.21 (1.17, 1.25) |
| Sociodemographic | Insurance Type - Medicare | 0.91 (0.87, 0.94) |
| Sociodemographic | Insurance Type - Other Payor type | 1.21 (1.15, 1.26) |
| Sociodemographic | Insurance Type - Uninsured | 1.73 (1.53, 1.95) |
| Sociodemographic | Race/Ethnicity - Asian | 1.78 (1.69, 1.89) |
| Sociodemographic | Race/Ethnicity - Hispanic | 3.40 (2.99, 3.86) |
| Sociodemographic | Race/Ethnicity - Non-Hispanic black | 3.06 (2.96, 3.16) |
| Sociodemographic | Sex - Male | 1.21 (1.19, 1.24) |
| Clinical/Setting | Abdominal Pain | 0.62 (0.59, 0.66) |
| Clinical/Setting | Acute Respiratory Distress Syndrome | 5.94 (5.27, |

| | | |
|---|---|---|
| | | 6.70) |
| Clinical/Setting | Acute Respiratory Failure | 1.74 (1.65, 1.83) |
| Clinical/Setting | Chest Pain | 0.62 (0.59, 0.66) |
| Clinical/Setting | Chills | 1.11 (1.01, 1.22) |
| Clinical/Setting | Confusion or Disorientation | 0.76 (0.71, 0.82) |
| Clinical/Setting | Congestion | 1.10 (1.01, 1.20) |
| Clinical/Setting | Conjunctivitis | 0.51 (0.39, 0.68) |
| Clinical/Setting | Cough | 1.45 (1.41, 1.49) |
| Clinical/Setting | Dehydration | 1.20 (1.12, 1.28) |
| Clinical/Setting | Disturbances of Smell or Taste | 7.31 (6.67, 8.01) |
| Clinical/Setting | Dizziness | 0.78 (0.71, 0.85) |
| Clinical/Setting | Dyspnea | 0.78 (0.75, 0.80) |
| Clinical/Setting | Edema | 0.42 (0.34, 0.51) |
| Clinical/Setting | Embolism and Thrombosis | 0.71 (0.65, 0.79) |
| Clinical/Setting | Fever | 2.05 (1.99, 2.11) |
| Clinical/Setting | GERD | 0.78 (0.74, 0.82) |
| Clinical/Setting | Headache | 1.10 (1.04, 1.17) |
| Clinical/Setting | Heart Failure | 0.53 (0.50, 0.57) |
| Clinical/Setting | Hemorrhage | 0.61 (0.52, 0.71) |
| Clinical/Setting | Hypoxia | 1.18 (1.13, 1.23) |
| Clinical/Setting | Index Setting - Inpatient | 1.14 (1.10, 1.17) |
| Clinical/Setting | Index Setting - Lab | 1.14 (1.09, 1.18) |
| Clinical/Setting | Index Setting - Other | 1.44 (1.39, 1.49) |
| Clinical/Setting | Loss of Appetite | 1.71 (1.48, 1.97) |

| | | |
|---|---|---|
| Clinical/Setting | Lower Respiratory | 1.86 (1.69, 2.05) |
| Clinical/Setting | Myalgia | 1.25 (1.17, 1.34) |
| Clinical/Setting | Number of positively-associated symptoms - 1-3 | 1.00 (0.96, 1.03) |
| Clinical/Setting | Number of positively-associated symptoms - 4-7 | 1.03 (0.95, 1.11) |
| Clinical/Setting | Number of positively-associated symptoms - 8-11 | 1.29 (0.89, 1.87) |
| Clinical/Setting | Other General Signs or Symptoms | 1.23 (1.19, 1.27) |
| Clinical/Setting | Other Specified Respiratory Disorders | 2.75 (2.58, 2.93) |
| Clinical/Setting | Pneumonia | 4.21 (4.04, 4.38) |
| Clinical/Setting | Positive for any seasonal respiratory isolate - At least 1 test positive | 0.26 (0.14, 0.46) |
| Clinical/Setting | Positive for any seasonal respiratory isolate - Tested but no positives | 0.85 (0.82, 0.87) |
| Clinical/Setting | Pre-Procedural Codes | 0.12 (0.11, 0.13) |
| Clinical/Setting | Rash | 0.46 (0.37, 0.56) |
| Clinical/Setting | Screening and Exposure | 0.66 (0.64, 0.67) |
| Clinical/Setting | Sepsis | 0.70 (0.67, 0.74) |
| Clinical/Setting | Stroke | 0.72 (0.66, 0.78) |
| Clinical/Setting | Tachycardia | 0.84 (0.82, 0.86) |
| Clinical/Setting | Thrombocytopenia | 1.22 (1.12, 1.32) |
| Clinical/Setting | Upper Respiratory Infection | 1.05 (1.01, 1.09) |
| Clinical/Setting | Vomiting | 0.74 (0.69, 0.79) |
| Comorbidities | Asthma | 0.93 (0.88, 0.97) |
| Comorbidities | BMI - obese | 1.34 (1.29, 1.40) |
| Comorbidities | BMI - overweight | 1.19 (1.14, 1.24) |
| Comorbidities | BMI - underweight | 0.81 (0.74, 0.89) |
| Comorbidities | Cancer | 0.81 (0.77, |

| | | |
|---|---|---|
| | | 0.85) |
| Comorbidities | Cerebrovascular disease | 1.13 (1.06, 1.20) |
| Comorbidities | Charlson Comorbidity Score - 1 | 0.77 (0.73, 0.80) |
| Comorbidities | Charlson Comorbidity Score - 2 | 0.58 (0.54, 0.63) |
| Comorbidities | Charlson Comorbidity Score - 3 | 0.48 (0.44, 0.54) |
| Comorbidities | Charlson Comorbidity Score - 4 | 0.40 (0.35, 0.45) |
| Comorbidities | Charlson Comorbidity Score - 5 | 0.33 (0.29, 0.39) |
| Comorbidities | Charlson Comorbidity Score - 6 | 0.27 (0.22, 0.32) |
| Comorbidities | Charlson Comorbidity Score - 7+ | 0.22 (0.17, 0.27) |
| Comorbidities | Chronic pulmonary disease | 0.89 (0.85, 0.94) |
| Comorbidities | Congestive heart failure | 1.19 (1.12, 1.27) |
| Comorbidities | Dementia | 2.66 (2.52, 2.82) |
| Comorbidities | Diabetes | 1.59 (1.51, 1.67) |
| Comorbidities | Hemiplegia or paraplegia | 1.14 (1.04, 1.25) |
| Comorbidities | Hyperlipidemia | 1.00 (0.97, 1.02) |
| Comorbidities | Hypertension | 0.92 (0.90, 0.95) |
| Comorbidities | Liver | 0.91 (0.86, 0.97) |
| Comorbidities | Peripheral vascular disease | 1.15 (1.09, 1.22) |
| Comorbidities | Pregnancy | 0.65 (0.61, 0.70) |
| Comorbidities | Renal disease | 1.19 (1.13, 1.25) |
| Comorbidities | Rheumatic disease | 0.89 (0.83, 0.96) |
| Comorbidities | Smoking status - Current smoker | 0.38 (0.36, 0.39) |
| Comorbidities | Smoking status - Previously smoked | 0.82 (0.80, 0.84) |
| Region and Study Week | Region - Northeast | 2.26 (1.97, 2.59) |

| | | |
|---|---|---|
| Region and Study Week | Region - South | 1.22 (0.98, 1.53) |
| Region and Study Week | Region - West | 0.39 (0.30, 0.51) |
| Region and Study Week | Weeks into study - [04,08) | 2.35 (2.14, 2.59) |
| Region and Study Week | Weeks into study - [08,12) | 1.47 (1.33, 1.62) |
| Region and Study Week | Weeks into study - [12,16) | 0.87 (0.78, 0.96) |
| Region and Study Week | Weeks into study - [16,20) | 0.89 (0.80, 0.98) |
| Region and Study Week | Weeks into study - [04,08) : Region - Northeast | 0.85 (0.74, 0.98) |
| Region and Study Week | Weeks into study - [08,12) : Region - Northeast | 0.72 (0.62, 0.83) |
| Region and Study Week | Weeks into study - [12,16) : Region - Northeast | 0.54 (0.47, 0.63) |
| Region and Study Week | Weeks into study - [16,20) : Region - Northeast | 0.23 (0.20, 0.27) |
| Region and Study Week | Weeks into study - [04,08) : Region - South | 0.40 (0.32, 0.51) |
| Region and Study Week | Weeks into study - [08,12) : Region - South | 0.39 (0.31, 0.50) |
| Region and Study Week | Weeks into study - [12,16) : Region - South | 0.60 (0.47, 0.77) |
| Region and Study Week | Weeks into study - [16,20) : Region - South | 1.27 (1.01, 1.60) |
| Region and Study Week | Weeks into study - [04,08) : Region - West | 1.14 (0.87, 1.51) |
| Region and Study Week | Weeks into study - [08,12) : Region - West | 0.88 (0.66, 1.17) |
| Region and Study Week | Weeks into study - [12,16) : Region - West | 0.74 (0.54, 1.01) |
| Region and Study Week | Weeks into study - [16,20) : Region - West | 1.64 (1.24, 2.19) |

* Population limited to patients with >365 days of database history in C2 with a definite result. Multivariate imputation by chained equations used to address missing values. Variables are mutually-adjusted.

Supplement M. Cohort 3: Odds Ratios and 95% Confidence Intervals Obtained from Multivariable Logistic Models (Corresponds to Forest Plot in Figure 5)

OR and 95% CI Cohort 3

| **Type** | **Term** | **OR (95% CI)** |
|---|---|---|
| Intercept | (Intercept) | 0.03 ( 0.03, 0.04) |
| Sociodemographic | Age - <18 | 1.90 ( 1.62, 2.23) |
| Sociodemographic | Age - 35-44 | 0.82 ( 0.74, 0.90) |
| Sociodemographic | Age - 45-54 | 1.01 ( 0.93, 1.10) |
| Sociodemographic | Age - 55-64 | 0.92 ( 0.85, 1.00) |
| Sociodemographic | Age - 65+ | 0.88 ( 0.79, 0.97) |
| Sociodemographic | Insurance Type - Medicaid | 1.91 ( 1.75, 2.08) |
| Sociodemographic | Insurance Type - Medicare | 0.94 ( 0.86, 1.03) |
| Sociodemographic | Insurance Type - Other Payor type | 1.22 ( 1.03, 1.43) |
| Sociodemographic | Insurance Type - Uninsured | 1.73 ( 0.38, 7.82) |
| Sociodemographic | Race/Ethnicity - Asian | 1.07 ( 0.95, 1.20) |
| Sociodemographic | Race/Ethnicity - Hispanic | 1.90 ( 1.63, 2.21) |
| Sociodemographic | Race/Ethnicity - Non-Hispanic black | 2.54 ( 2.32, 2.77) |
| Sociodemographic | Sex - Male | 1.16 ( 1.11, 1.22) |
| Clinical/Setting | Days since COVID-like illness - 0 | 1.97 ( 1.81, 2.14) |
| Clinical/Setting | Days since COVID-like illness - 1 - 3 | 1.96 ( 1.71, 2.25) |
| Clinical/Setting | Days since COVID-like illness - 4 - 7 | 2.06 ( 1.74, 2.42) |
| Clinical/Setting | Days since COVID-like illness - 8 - 14 | 2.40 ( 2.04, 2.83) |
| Clinical/Setting | Days since COVID-like illness - 15 - 21 | 2.21 ( 1.83, 2.68) |
| Clinical/Setting | Days since COVID-like illness - 22 - 28 | 3.20 ( 2.66, 3.84) |
| Clinical/Setting | Days since COVID-like illness - 29 - 35 | 3.67 ( 3.12, 4.31) |
| Clinical/Setting | Days since COVID-like illness - 36 - 42 | 4.53 ( 3.97, 5.18) |

| | | |
|---|---|---|
| Clinical/Setting | Index Setting - Inpatient | 0.73 ( 0.57, 0.92) |
| Clinical/Setting | Index Setting - Lab | 1.06 ( 0.99, 1.15) |
| Clinical/Setting | Index Setting - Other | 1.08 ( 1.00, 1.16) |
| Clinical/Setting | Max previous viral test results (positive > negative > unknown) - Negative | 0.54 ( 0.49, 0.58) |
| Clinical/Setting | Max previous viral test results (positive > negative > unknown) - Non-definitive result | 3.21 ( 1.89, 5.46) |
| Clinical/Setting | Max previous viral test results (positive > negative > unknown) - Positive | 44.16 (37.26, 52.33) |
| Clinical/Setting | Other General Signs Or Symptoms | 1.33 ( 1.17, 1.50) |
| Clinical/Setting | Routine Exam | 0.88 ( 0.82, 0.94) |
| Clinical/Setting | Screening and procedure codes combined | 0.99 ( 0.94, 1.04) |
| Comorbidities | AIDS/HIV | 0.76 ( 0.60, 0.97) |
| Comorbidities | BMI - obese | 1.36 ( 1.26, 1.47) |
| Comorbidities | BMI - overweight | 1.21 ( 1.12, 1.30) |
| Comorbidities | BMI - underweight | 0.80 ( 0.65, 0.98) |
| Comorbidities | Charlson Comorbidity Score - 1 | 0.90 ( 0.84, 0.95) |
| Comorbidities | Charlson Comorbidity Score - 2 | 0.76 ( 0.69, 0.84) |
| Comorbidities | Charlson Comorbidity Score - 3 | 0.73 ( 0.64, 0.83) |
| Comorbidities | Charlson Comorbidity Score - 4 | 0.80 ( 0.67, 0.96) |
| Comorbidities | Charlson Comorbidity Score - 5 | 0.55 ( 0.43, 0.72) |
| Comorbidities | Charlson Comorbidity Score - 6 | 0.56 ( 0.39, 0.80) |
| Comorbidities | Charlson Comorbidity Score - 7+ | 0.52 ( 0.34, 0.80) |
| Comorbidities | Dementia | 1.46 ( 1.07, 1.99) |
| Comorbidities | Diabetes | 1.38 ( 1.27, 1.50) |
| Comorbidities | Hemiplegia or paraplegia | 1.44 ( 0.97, 2.13) |
| Comorbidities | Smoking status - Current smoker | 0.51 ( 0.45, |

| | | |
|---|---|---|
| | | 0.58) |
| Comorbidities | Smoking status - Previously smoked | 0.86 ( 0.81, 0.91) |
| Region and Study Week | Region - Northeast | 4.53 ( 4.22, 4.86) |
| Region and Study Week | Region - South | 0.90 ( 0.75, 1.07) |
| Region and Study Week | Region - West | 1.00 ( 0.76, 1.32) |
| Region and Study Week | Weeks into study cat (antibody) - [04,08) | 0.92 ( 0.77, 1.11) |
| Region and Study Week | Weeks into study cat (antibody) - [08,12) | 0.75 ( 0.63, 0.90) |
| Region and Study Week | Weeks into study cat (antibody) - [12,16) | 0.74 ( 0.61, 0.89) |

* Population limited to patients with >365 days of database history in C3 with a definite result. Multivariate imputation by chained equations used to address missing values. Variables are mutually-adjusted.
